# Microbial signatures in the lower airways of mechanically ventilated COVID19 patients associated with poor clinical outcome

**DOI:** 10.1101/2021.02.23.21252221

**Authors:** Imran Sulaiman, Matthew Chung, Luis Angel, Jun-Chieh J. Tsay, Benjamin G. Wu, Stephen T. Yeung, Kelsey Krolikowski, Yonghua Li, Ralf Duerr, Rosemary Schluger, Sara A. Thannickal, Akiko Koide, Samaan Rafeq, Clea Barnett, Radu Postelnicu, Chang Wang, Stephanie Banakis, Lizzette Perez-Perez, George Jour, Guomiao Shen, Peter Meyn, Joseph Carpenito, Xiuxiu Liu, Kun Ji, Destiny Collazo, Anthony Labarbiera, Nancy Amoroso, Shari Brosnahan, Vikramjit Mukherjee, David Kaufman, Jan Bakker, Anthony Lubinsky, Deepak Pradhan, Daniel H. Sterman, Michael Weiden, Adriana Hegu, Laura Evans, Timothy M. Uyeki, Jose C. Clemente, Emmie De wit, Ann Marie Schmidt, Bo Shopsin, Ludovic Desvignes, Chan Wang, Huilin Li, Bin Zhang, Christian V. Forst, Shohei Koide, Kenneth A. Stapleford, Kamal M. Khanna, Elodie Ghedin, Leopoldo N. Segal

## Abstract

Mortality among patients with COVID-19 and respiratory failure is high and there are no known lower airway biomarkers that predict clinical outcome. We investigated whether bacterial respiratory infections and viral load were associated with poor clinical outcome and host immune tone. We obtained bacterial and fungal culture data from 589 critically ill subjects with COVID-19 requiring mechanical ventilation. On a subset of the subjects that underwent bronchoscopy, we also quantified SARS-CoV-2 viral load, analyzed the microbiome of the lower airways by metagenome and metatranscriptome analyses and profiled the host immune response. We found that isolation of a hospital-acquired respiratory pathogen was not associated with fatal outcome. However, poor clinical outcome was associated with enrichment of the lower airway microbiota with an oral commensal (*Mycoplasma salivarium*), while high SARS-CoV-2 viral burden, poor anti-SARS-CoV-2 antibody response, together with a unique host transcriptome profile of the lower airways were most predictive of mortality. Collectively, these data support the hypothesis that 1) the extent of viral infectivity drives mortality in severe COVID-19, and therefore 2) clinical management strategies targeting viral replication and host responses to SARS-CoV-2 should be prioritized.

## Introduction

The earliest known case of Severe Acute Respiratory Syndrome Coronavirus 2 (SARS-CoV-2) infection causing coronavirus virus disease (COVID-19) is thought to have occurred on November 17, 2019^1^. As of February 20, 2021, 110.3 million confirmed cases of COVID-19 and 2.4 million deaths have been reported worldwide^2^. As the global scientific community rallied in a concerted effort to understand SARS-CoV-2 infections, our background knowledge is rooted in previous experience with the related zoonotic betacoronaviruses Middle East Respiratory Syndrome coronavirus (MERS-CoV) and SARS-CoV-1 that have caused severe pneumonia with 34.4% and 9% case fatality, respectively^3^. As observed for these related coronaviruses, SARS-CoV-2 infection can result in an uncontrolled inflammatory response^4^ leading to acute respiratory distress syndrome (ARDS) and multi-organ failure, both associated with increased mortality. While a large proportion of the SARS-CoV-2 infected population is asymptomatic or experiences mild illness, a substantial number of individuals will develop severe disease and require hospitalization, with some progressing to respiratory failure. Mortality among hospitalized COVID-19 patients is estimated to be approximately 20%, which can go up to 70% among those requiring invasive mechanical ventilation ^5–12^.

Mortality in other viral pandemics, such as the 1918 H1N1 and 2009 H1N1 influenza pandemics, has been attributed in part to bacterial co-infection or super-infection ^13,14^. To determine if this is also the case for COVID-19, we can use next generation sequencing (NGS) to probe the complexity of the microbial environment (including RNA and DNA viruses, bacteria and fungi) and how the host (human) responds to infection.

Recent studies have used this approach to uncover microbial signatures in patients with ARDS.^15,16^ Increased bacterial burden and the presence of gut-associated bacteria in the lung were shown to worsen outcomes in these critically ill patients ^15,17^, highlighting the potential role of the lung microbiome in predicting outcomes in ARDS. In a recent study using whole genome sequencing to profile the gut microbiome of 69 patients from Hong Kong, investigators identified an increased abundance of opportunistic fungal pathogens among patients with confirmed COVID-19^18^. While there is emerging interest in understanding the microbial environment in patients with SARS-CoV-2 infections, few studies have attempted to characterize this at the primary site of the disease activity: the lower airways^19,20^. Furthermore, no study has yet determined whether microbial differences in the airways of COVID-19 patients could be contributing to the different outcomes in patients receiving mechanical ventilation.

In this investigation, we accessed a large prospective cohort of critically ill patients with SARS-CoV-2 infection who required invasive mechanical ventilation, and from whom bronchoalveolar lavage (BAL) samples were collected. We characterized the lung microbiome of these patients in parallel with analyses of lower airway markers of host immunity. While we did not find that isolation of a secondary respiratory pathogen was associated with prolonged mechanical ventilation (>28 days) or fatal outcome, we did identify critical microbial signatures—characterized by enrichment of oral commensals, high SARS-CoV-2 load, and decreased anti-SARS-CoV-2 IgG response—associated with fatal outcome, suggesting a need for more targeted antiviral therapeutic approaches for the care of critically ill COVID19 patients.

## Results

### Cohort

From March 3^rd^ to June 18^th^ 2020, a total of 589 patients with laboratory-confirmed SARS-CoV-2 infection were admitted to the intensive care units of two academic medical centers of NYU Langone Health in New York (Long Island and Manhattan) and required invasive mechanical ventilation (MV). This included a subset of 142 patients from the Manhattan campus who underwent bronchoscopy for airway clearance and/or tracheostomy from which we collected and processed lower airway samples for this investigation (**Supplementary Fig. 1**). **Table 1** shows demographics and clinical characteristics of the 142 patients who underwent bronchoscopy divided into three clinical outcomes: survivors with ≤28 Days on MV; survivors with >28 Days on MV; and deceased. The median post admission follow-up time was 232 days (CI=226-237 days). **Supplementary Tables 1 and 2** compare similar data across all 589 subjects, divided per site and sub-cohorts. Patients at the Manhattan campus who underwent bronchoscopy were younger, had lower body mass index (BMI), and a lower prevalence of chronic obstructive pulmonary disease (COPD; **Supplementary Table 1**). Among the cohort that provided lower airway samples through bronchoscopy, 37% of the subjects were successfully weaned within 28 days of initiation of MV and survived hospitalization, 39% required prolonged MV but survived hospitalization, and 23% died. Patients within the bronchoscopy cohort had a higher overall survival than the rest of the NYU COVID-19 cohort since most critically ill patients were not eligible for bronchoscopy or tracheostomy. Mortality among those in the no-bronchoscopy cohort was 77%. In the overall NYU cohort, higher age and BMI were associated with increased mortality (**Supplementary Table 2**). There was a similar, albeit non-significant, trend for the bronchoscopy cohort. Among the clinical characteristics of this cohort, patients within the deceased group more commonly had a past medical history of chronic kidney disease and cerebrovascular accident.

Study patients were admitted during the first wave of the pandemic in the US, prior to current standardized management of COVID-19. Within the bronchoscopy cohort, more than 90% of the subjects received hydroxychloroquine and anticoagulation (therapeutic dose), 69% received corticosteroids, 41% received tocilizumab (anti-Interleukin (IL)-6 receptor monoclonal antibody), 21% required dialysis, and 18.9% were started on extracorporeal membrane oxygenation (ECMO) (**Table 1**). Antimicrobial therapy included use of antivirals (lopinavir/ritonavir in 16% and remdesivir in 10%), antifungals (fluconazole in 40% and micafungin in 57%), and antibiotics (any, in 90% of the subjects). Among the factors associated with clinical outcome within the bronchoscopy cohort, patients who survived were more commonly placed on ECMO whereas patients who died had frequently required dialysis (**Table 1**); these trends were also observed across the whole NYU cohort. Neither hydroxychloroquine or azithromycin were significantly associated with clinical outcome; however, patients who survived were more frequently treated with the combination antibiotic piperacillin/tazobactam.

Within the first 48hrs from admission, respiratory bacterial cultures were rarely obtained (n=70/589, 12%) with very few positive results (n=12, 17%). Blood cultures were more commonly obtained (n=353/589, 60%) but the rate of bacterial culture positivity was much lower (n=5, 1.4%). These data support that community acquired bacterial co-infection was not a common presentation among critically ill COVID-19 patients.

During their hospitalization, most patients had respiratory and/or blood specimens collected for bacterial cultures (**Table 1** and **Supplementary Table 1**). The proportions of positive bacterial respiratory cultures and blood cultures were 73% and 43%, respectively. We evaluated whether respiratory or blood culture results obtained as per clinical standard of care were associated with clinical outcome. Risk analyses for the culture results during hospitalization for the whole cohort (n=589) demonstrated that bacterial culture positivity was not associated with increased odds of dying but was associated with prolonged mechanical ventilation in the surviving patients (**Figure 1**). Since length of stay could potentially affect these results (patients who died could have a shorter hospitalization, and therefore may have had fewer specimens collected for cultures), we repeated the analysis using culture data obtained during the first two weeks of hospitalization. This analysis showed that bacterial pathogen culture positivity (both respiratory and blood) during the early period of hospitalization was not associated with worse outcome (**Figure 1** and **Supplementary Table 3**). Interestingly, identification of oral bacteria in respiratory culture, commonly regarded as procedural contaminants, was associated with higher odds of prolonged mechanical ventilation (>28 days) among survivors. Similar trends were noted when analysis was performed on subjects from NYU LI and NYU Manhattan separately, or for the bronchoscopy cohort (**Supplementary Table 2**). Among the bronchoscopy cohort, there was no statistically significant association between culture results and clinical outcome, but there was a trend towards an increased rate of positive respiratory cultures for *Staphylococcus aureus* (including MRSA), *S. epidermidis*, and *Klebsiella pneumoniae* in the survival groups (**Table 1**). These data suggest that in critically ill patients with COVID-19 requiring MV, hospital isolation of a secondary respiratory bacterial pathogen is not associated with worse clinical outcome.

**Figure 1.**
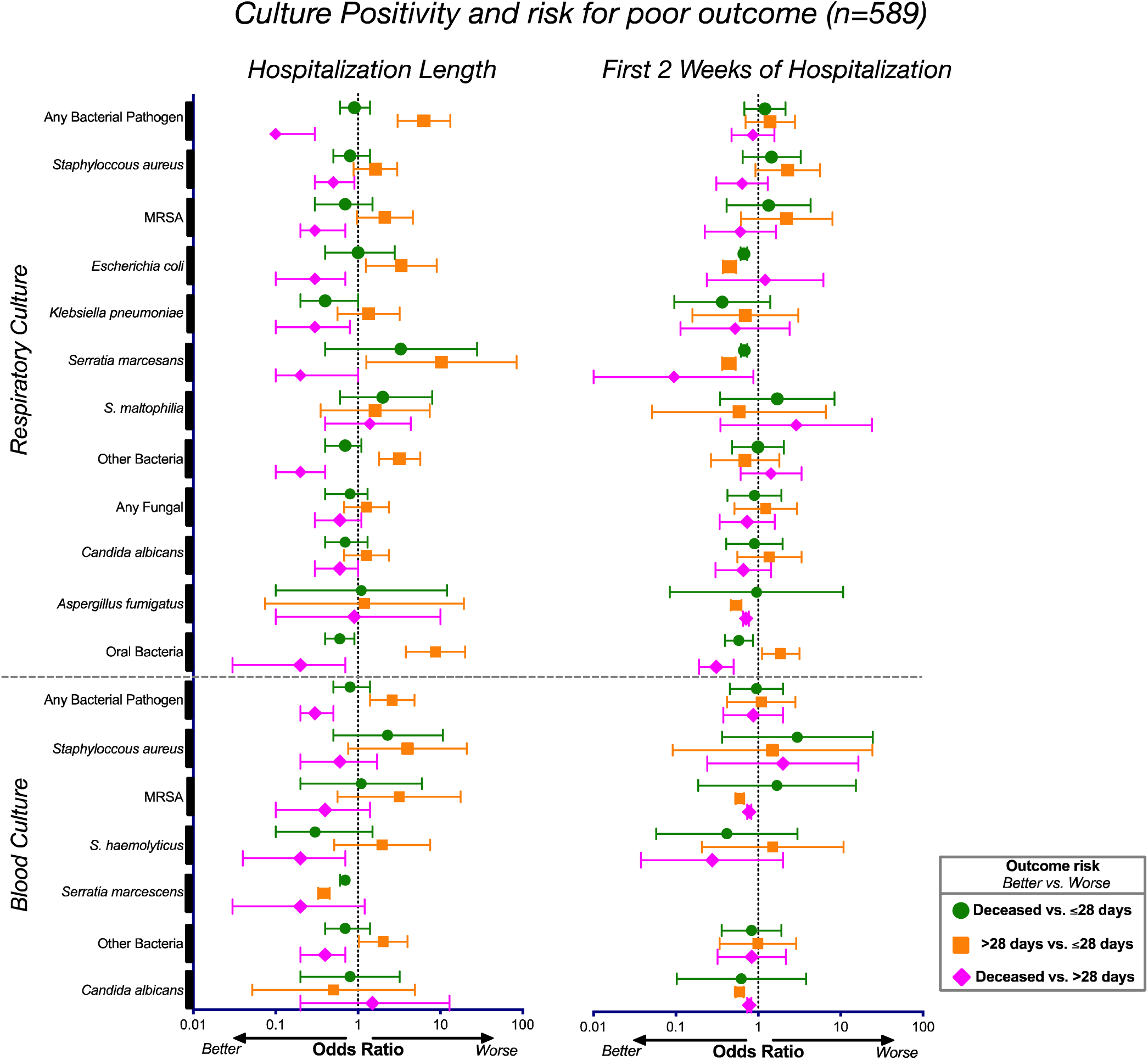
Associations between culture positivity and clinical outcome. Odds ratios and corresponding 95% confidence intervals for rates of culture positivity for the whole cohort (n=589) during the length of their hospitalization (left) and during the first 2 weeks of hospitalization (right).

### SARS-CoV-2 load in the lower airways is associated with poor clinical outcome

Using supraglottic and BAL samples from patients undergoing bronchoscopy (n=142), we evaluated the viral load by rRT-PCR for the SARS-CoV-2 N gene, adjusted by levels of human ribosomal protein (RP). Of note, the majority of samples were largely obtained in the second week of hospitalization (**Table 1**, median[IQR] = 10[6-14], 13[8-16], and 13[8-16] for the ≤28-days MV, >28-days MV, and deceased groups, respectively, p=ns). Paired analysis of upper and lower airway samples revealed that, while there was a positive association between SARS-CoV-2 viral load of the paired samples (rho = 0.60, p<0.0001), there was a subset of subjects (21%) for which the viral load was greater in the BAL than in the supraglottic area, indicating topographical differences in SARS-CoV-2 replication (**Figure 2a**). Importantly, while the SARS-CoV-2 viral load in the upper airway samples was not associated with clinical outcome (**Supplementary Fig. 2**), patients who died had higher viral load in their lower airways than patients who survived (**Figure 2b**). We then evaluated virus viability in BAL samples by measuring levels of subgenomic RNA (sgRNA) targeting the E gene of SARS-CoV-2. This mRNA is only transcribed inside infected mammalian cells and is not packed into virions, thus, its presence is indicative of viable infecting viral particles in a sample^21^. In BAL, levels of sgRNA correlated with viral load as estimated by rRT-PCR for the SARS-CoV-2 N gene (**Figure 2c**) and the highest percentage of measurable sgRNA was in the deceased group followed by the ≤28-days MV group, and the >28-days MV group (17,7%, 11.5%, and 3.7%, respectively, chi-square p=0.028 for the comparison deceased vs. >28-days MV group). Thus, while in most cases levels of sgRNA were not measurable in BAL suggesting that there were no viable viral particles in the lower airways of COVID-19 patients at the time of bronchoscopy (overall median[IQR] = 12[7-16] days from hospitalization), the lower airway viral burden, as estimated by rRT-PCR, is associated with mortality in critically ill COVID-19 patients.

**Figure 2.**
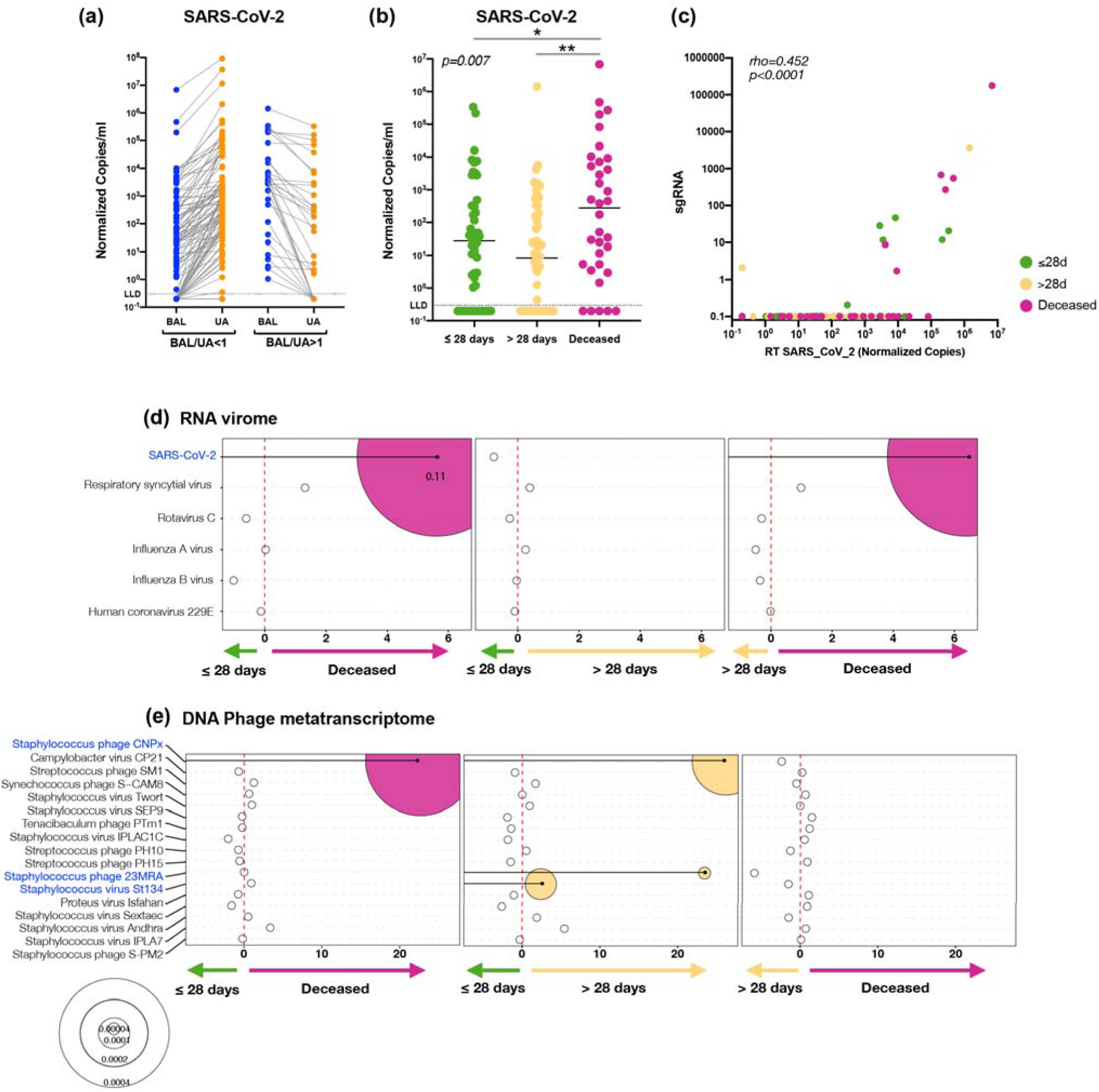
SARS-CoV-2 viral load and virus metatranscriptome analyses. Copies of the SARS-CoV-2 N gene per ml, normalized by the Human RNase P gene, comparing paired upper and lower airway samples **(a)** and levels in BAL comparing clinical outcome groups **(b**, *= Mann–Whitney U p<0.05, **= Mann–Whitney U p<0.01). **(c)** PCoA analysis based on Bray Curtis Dissimilarity index of BAL Metatranscriptome data comparing clinical outcome (PERMANOVA p-value). Bubble plot showing DESeq results of RNA viruses **(d)** and expressed DNA phages **(e)** enriched in each clinical outcome comparisons (bubble size based on median relative abundance for those found statistically significant).

### Microbial community structure of the lower airways is distinct from the upper airways in critically ill patients

Considering the bacterial species and the viral loads identified in the lower and upper airways of this cohort and their association with outcomes, we profiled in detail their viral and microbial composition. Microbial communities were evaluated using parallel datasets of RNA and DNA sequencing from 118 COVID-19 patients with lower airway samples that passed appropriate quality control and a subset of paired 64 upper airway samples, along with background bronchoscope controls.

RNA sequencing (RNAseq) of the metatranscriptome provided insight into the RNA virome as well as the transcriptomes of DNA viruses, bacteria, and fungi. Given the low biomass of lower airway samples we first identified taxa as probable contaminants by comparing the relative abundance between background bronchoscope and BAL samples (**Supplementary Fig. 3a** and **Supplementary Table 4**). However, we did not remove any taxa identified as probable contaminants from subsequent analyses. A comparison of the microbial community complexity captured in these data, determined using the Shannon diversity Index, showed there was significantly lower α diversity in the lower airway samples than in the upper airways and background controls (**Supplementary Fig. 4a**). Similarly, β diversity analysis based on the Bray Curtis Dissimilarity index indicated that the microbial composition of the lower airways was distinct from the upper airways and background controls (**Supplementary Fig. 4b**, PERMANOVA p<0.01). Sequence reads indicated a much higher relative abundance of SARS-CoV-2 in the lower than in the upper airways for this cohort (**Supplementary Fig. 4c)**. Comparisons of the most dominant bacterial and fungal taxa that were functionally active showed that *S. epidermidis*, *Mycoplasma salivarium, S. aureus*, *Prevotella oris*, and *Candida albicans,* many often-considered oral commensals, were present in both upper and lower airway samples (**Supplementary Fig. 4c**). Interestingly, the lytic phage Proteus virus Isfahan, known to be active against biofilms of *Proteus mirabilis* ^22^, was found to be highly transcriptionally active in the BAL.

DNA sequencing data provided insight into the DNA virome, as well as the bacterial and fungal metagenomes. As for the metatranscriptome data, we first identified taxa as probable contaminants but these were not removed for subsequent analyses (**Supplementary Fig. 3b**). Both α and β diversity analyses of the metagenome support distinct microbial community features in the lower airways as compared with the upper airways and background controls (**Supplementary Fig. 5a, 5b**). Among the top 10 taxa across lower and upper airway samples were *S. aureus*, *Salmonella enterica*, *Burkholderia dolosa*, and *Klebsiella variicola*. *Candida albicans* only ranked #77 in the BAL while it was ranked 5^th^ in the metatranscriptome data indicating that while present at low relative abundance, it was highly active (**Supplementary Table 4**). *K. variicola*, while prevalent at a high relative abundance (#4 in BAL, and #5 in the upper airways) in patients of this cohort, its ranking in the RNAseq data was not among the top 50, indicating that it was not as active functionally as other bacteria. Conversely, while *S. epidermis* ranked as the most highly functional taxon in both lower and upper airways, based on RNAseq reads (**Supplemental Fig. 3c**), it was 33^rd^ in relative abundance in the BAL DNAseq data but was present at very high relative abundance in the upper airways (ranked #3). These data suggest that microbes that colonize the upper airways and the skin were common in the lower airways in this cohort of COVID-19 patients requiring invasive mechanical ventilation.

### Distinct microbial signatures are associated with different clinical outcomes

To determine the potential impact of vertebrate viruses on outcome, we compared virus enrichment differences in BAL samples across the three clinical outcome groups (≤28-days MV, >28-days MV, and deceased). As it pertains to the vertebrate RNA virome subfraction, there were significant differences (β diversity) between the three clinical outcome groups (**Supplementary Fig. 6**, PERMANOVA p<0.01). There were no significant differences for the vertebrate DNA virome or DNA virus transcriptome subfractions of the sequence reads (data not shown). Consistent with the SARS-CoV-2 viral load assessed by RT-PCR, differential expression analysis (DESeq) of the RNA virome identified SARS-CoV-2 as being enriched in the deceased group, as compared with both ≤28-days and >28-days MV groups (fold change >5, **Figure 2d**). Cox proportional hazards modeling supports that enrichment with SARS-CoV-2 was associated with increased risk for death (HR 1.33, 95% CI= 1.07-1.67, pvalue=0.011, FDR adjusted pvalue=0.06; **Supplementary Table 5**).

Analysis of differential DNA virus abundance using DEseq did not show statistically significant differences. Because the virome includes viruses of bacteria and archaea, we also analyzed the phage data (including viruses of archaea). Phages impact the bacterial population—including bacterial pathogens—and so could be clinically relevant. At a compositional level, the virome of DNA phages did not display statistically significant differences or significant virus enrichment based on clinical outcome groups (data not shown). However, while the phage metatranscriptome α and β diversity was similar across the clinical outcome groups, there were various taxonomic differences at the RNA level with enrichment of *Staphylococcus* phages CNPx in the deceased and >28-day MV groups when compared with the ≤28-day MV group (**Figure 2e**). Differential expression from two other *Staphylococcus* phages was also observed in the >28-days MV group as compared with the ≤28-days MV group (**Figure 2e**). None of the described taxa were identified as possible contaminants (**Supplementary Table 4**).

### Enrichment of the lower airway microbiota with oral commensals is associated with poor outcome

We evaluated the overall bacterial load by quantitative PCR, targeting the 16S rRNA gene. As expected, the bacterial load in the lower airways was several folds lower than in the upper airways but clearly higher than the background bronchoscope control (**Supplementary Fig. 7**). Patients who died had higher total bacterial load in their lower airways than patients who survived (**Figure 3a**).

**Figure 3.**
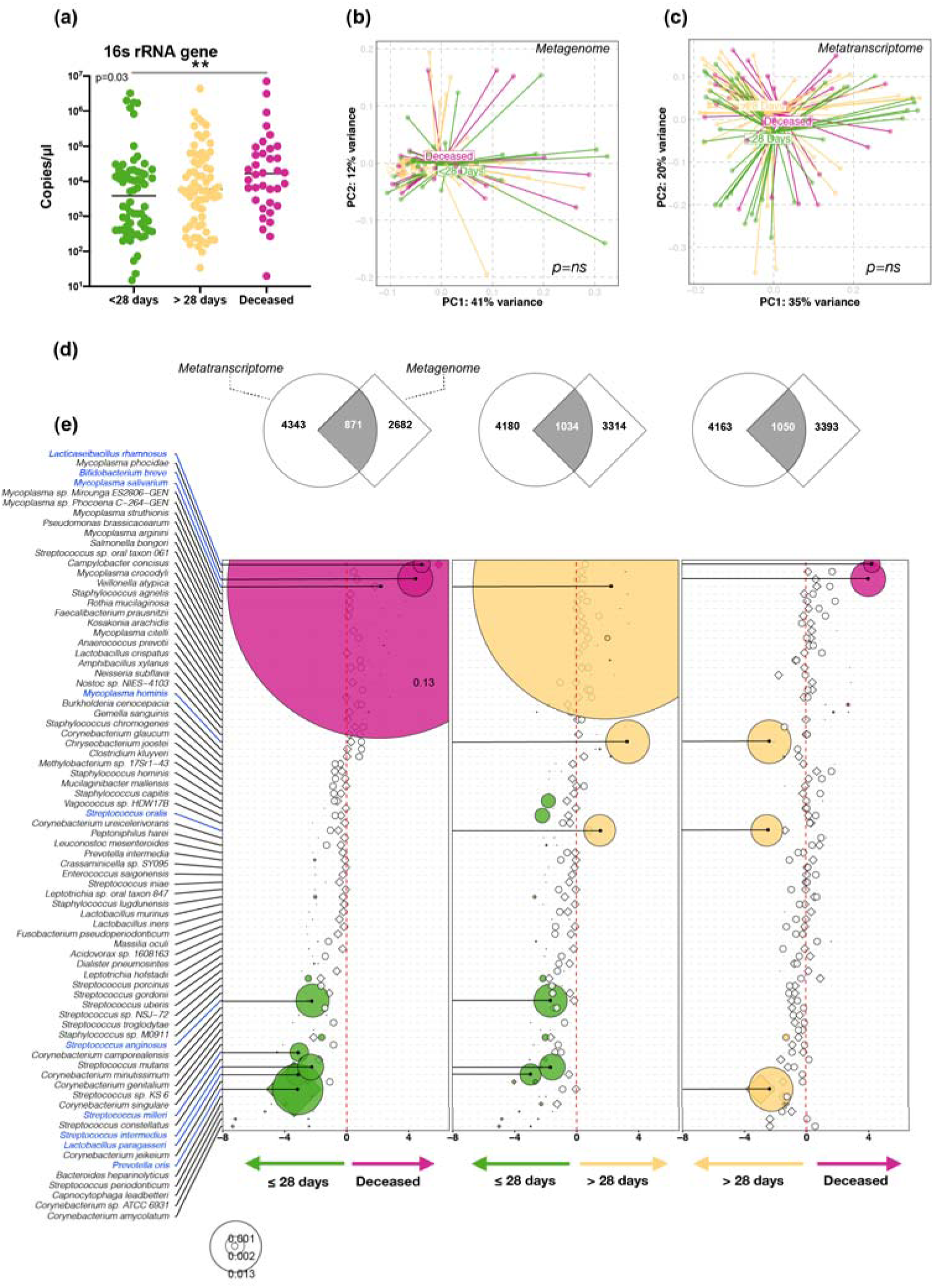
Bacteria load and taxonomic compositional analyses. **(a)** Bacterial load measured by ddPCR targeting 16S rRNA gene (**= Mann–Whitney U p<0.01). PCoA analysis based on Bray Curtis Dissimilarity index of BAL Metagenome **(b)** and Metatranscriptome **(c)** data comparing clinical outcome (PERMANOVA p-value). **(d)** Gene Set Enrichment Analysis (GSEA) was used to compare the taxonomic signatures identified in BAL metagenome (diamonds) and metatranscriptome (circles) as distinctly enriched for comparisons between clinical outcome groups (differential enrichment performed based on DESeq2 analysis). **(e)** Bubble plot showing DESeq results of bacteria found concordantly differentially enriched between clinical outcome groups (bubble size based on median relative abundance for those found statistically significant).

While no statistically significant differences were noted in α or β diversity across clinical outcome groups (**Figure 3b-c**), several differences were noted when differential enrichment was evaluated using DESeq. For the comparisons made across the clinical outcome groups we focused on consistent signatures identified in the lower airway metagenome and metatranscriptome. Coherence of differentially enriched taxa was determined by gene set enrichment analysis (GSEA) (**Figure 3d**) and directionality of enrichment between the two datasets was evaluated (**Figure 3e**). Among the most abundant taxa, the oral commensal *M. salivarum* was enriched in the deceased and >28-days MV groups as compared with the ≤28-days MV group. In contrast, a different oral commensal, *Prevotella oris*, was enriched in the ≤28-days MV group as compared with the deceased and >28-days MV groups. These data support that oral commensals are frequently found in the lower airways of critically ill COVID-19 patients and that differences between groups could be due to differential microbial pressures related to host factors. Interestingly, most of the statistically significant taxa were identified in the metatranscriptome rather than in the metagenome data, with only *P. oris* identified in both datasets. None of the described taxa were identified as possible contaminants (**Supplementary Table 4**). Overall, most of the microbial signatures identified as enriched in the deceased or in subjects on prolonged MV are regular colonizers of healthy skin and mucosal surfaces rather than frequent respiratory pathogens.

For the fungal data, there were no statistically significant differences in α or β diversity identified between clinical outcome groups in the metagenome or the metatranscriptome data (**Supplementary Fig. 8a** and **8c**). However, in the metagenome data, we identified *Candida glabrata* enriched in the deceased group as compared with the ≤28-days MV and the >28-days MV groups but this was not consistent in the metatranscriptome data (**Supplementary Fig. 8b** and **8d**).

### Poor clinical outcomes are associated with enrichment of antimicrobial resistance genes and glycosphingolipid biosynthesis

We used the gene annotation of the DNAseq and RNAseq data to profile the microbial functional potential of the lower airway samples. For the comparisons made across the clinical outcome groups, we focused on consistent functional signatures identified in the lower airway metagenome and metatranscriptome. Coherence of differentially enriched functions was determined using GSEA (**Figure 4a**) and directionality of enrichment was also evaluated (**Figure 4b**). Overall, there was coherence of directionality between the metranscriptomics and metagenomics datasets for the comparisons between deceased vs ≤28-days MV, and >28-days MV vs ≤28-days MV groups. Interestingly, statistically significant differences were only noted in the metatranscriptome data and not in the metagenome data. Among the top differentially expressed pathways in the poor outcome groups were glycosylases, oxidoreductase activity, transporters, and two-component system, among other genes. The two-component system is used by bacteria and fungi for signaling. A specific analysis of antibiotic resistance genes shows that there was significant gene enrichment and expression of biocide resistance in the deceased group as compared to the two other MV groups (**Supplementary Fig. 9).** There was also significant expression of genes resistant to trimethoprim and phenolic compound, as well as multi drug resistance in the deceased group as compared to the ≤28-days MV group. Presence of the resistance gene against Trimethropim was not significantly associated with prior exposure with Trimethoprim. However, only 7 patients received this drug before sample collection.

**Figure 4.**
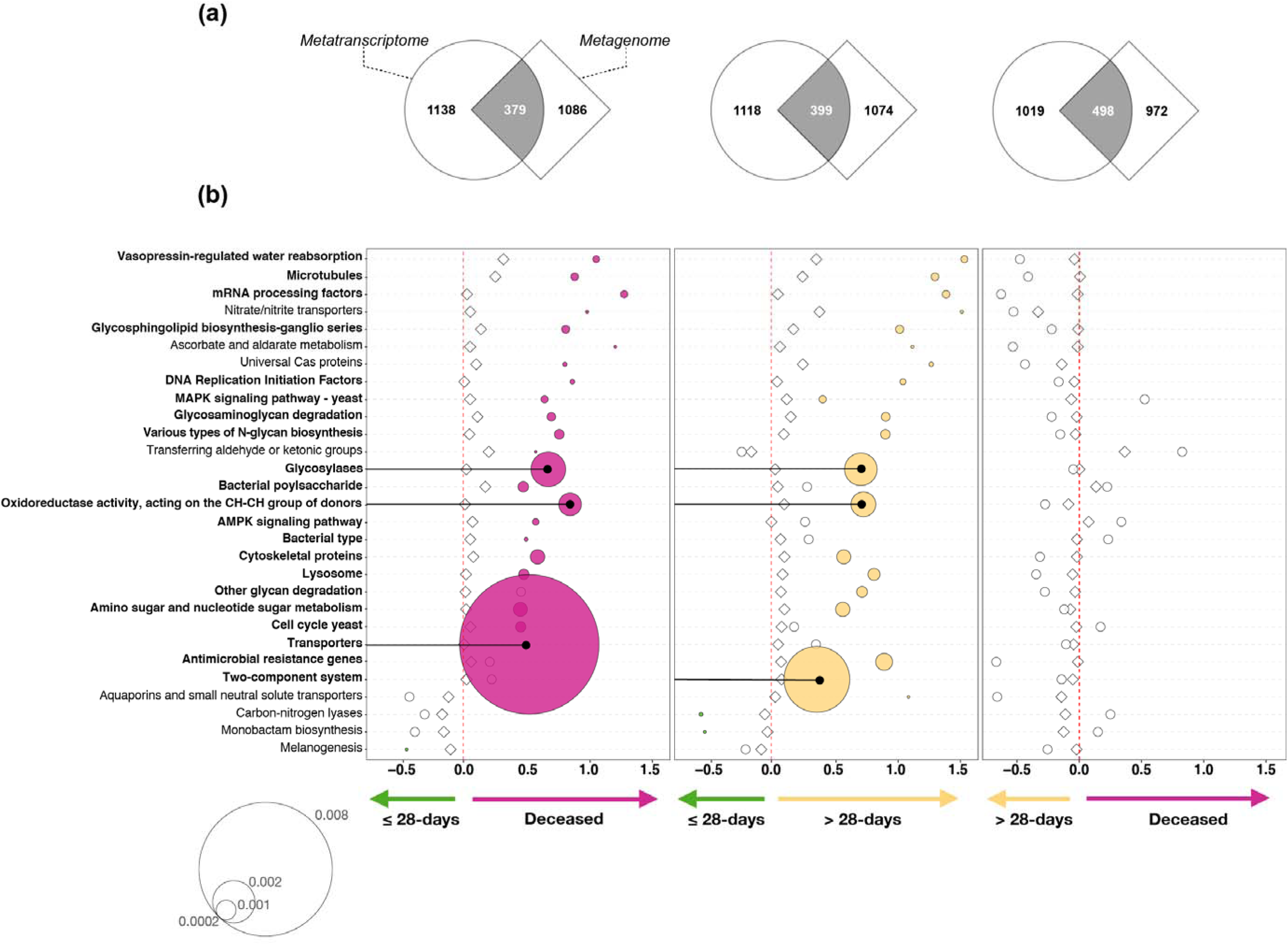
Functional microbial compositional analyses. KOs were summarized to associated pathways and differential expression was calculated based on DESeq2 analysis. **(a)** Gene Set Enrichment Analysis (GSEA) was used to compare the functional signatures identified in BAL metagenome and metatranscriptome as distinctly enriched for comparisons between clinical outcome groups. **(b)** Bubble plot showing DESeq results of microbial functions found concordantly differentially enriched between clinical outcome groups (bubble size based on median relative abundance for those found statistically significant).

### Lower airway host immune phenotype shows failure of adaptive and innate immune response to SARS-CoV-2 among deceased subjects

To evaluate the host immune response to SARS-CoV-2 infection, we first measured levels of anti-Spike and anti-RBD (receptor binding domain) antibodies in BAL samples. For both anti-Spike and anti-RBD immunoglobulins, levels of IgG, IgA and IgM were several logs higher than levels found in BAL samples from non-SARS-CoV-2 infected patients. Importantly, IgG levels of anti-Spike and anti-RBD were significantly lower in the deceased group as compared to the levels found in patients who survived (**Figure 5a** and **Supplementary Fig. 10a-c**, p<0.05). A neutralization assay performed using BAL fluid showed varying levels of neutralization across all samples (as estimated by EC50) but no statistically significant differences between the clinical outcome groups (**Supplementary Fig. 10d**). We then evaluated whether levels of antibodies correlated with viral load in BAL samples. While viral load levels of SARS-CoV-2 measured with rRT-PCR did not correlate with BAL measurements of SARS-CoV-2 specific antibodies, sgRNA viral load levels negatively correlated with BAL levels of Anti-Spike (IgG and IgA), Anti-RBD (IgG and IgA) and the Neutralization assay (**Supplementary Table 6**). These data suggest that the IgG subfraction is an important marker of the adaptive immune response in the lung of critically ill COVID-19 patients and that both sub-fractions of IgG and IgA anti-SARS-CoV-2 may contribute to the viral replication control in the lower airways.

Host transcriptome analyses of BAL samples showed significant differences across clinical outcome groups based on β diversity composition (**Supplementary Fig. 11)**. We identified multiple differentially expressed genes across the clinical outcome groups (**Supplementary Fig. 11b-d**). First, we noted that the lower airway transcriptomes showed downregulation of heavy constant of IgG (IGHG3), and heavy constant of IgA (IGHA1) genes in those with worse clinical outcome (**Supplementary Table 7**). We then used IPA (Ingenuity Pathway Analysis) to summarize differentially expressed genes across the three clinical outcome groups (**Figure 5b**). The sirtuin Signaling Pathway (a pathway known to be involved in aging, gluconeogenesis/lipogenesis, and host defense against viruses)^23^ and the ferroptosis pathway (an iron-dependent form of regulated cell death present in bronchial epithelium)^24,25^ were both upregulated in those with worse outcome. While this may reflect the host response to viral infection, other differences in the transcriptomic data showed downregulation of mitochondrial oxidative phosphorylation, HIF1α, STAT3, and Phospholipase C Signaling. Additional canonical signaling pathways, including insulin secretion, multiple Inositol related pathways, noradrenaline/adrenaline degradation signaling, and xenobiotic related metabolism were significantly downregulated when comparing the >28-days MV vs. ≤28-days MV groups. Upstream pathway prediction analyses of the host airway transcriptome support previously reported mitochondria dysfunction^26^ (inhibition in mitochondrial related regulators NSUN3, MRPL14, MRPL12, LONP1, DAP3), and metabolic/gluconeogenesis dysregulation^27,28^ (SIRT3) in critically ill COVID-19 subjects with poor outcome **(Supplementary Table 8)**. We also observed decreased activation in the inflammatory response in critically ill COVID-19 subjects with poor outcome (phagocytes, neutrophils, and granulocytes, and leukocytes**; Supplementary Table 9**). A comparison of clinical outcome between the >28-days MV vs. ≤28-days MV groups showed upstream predicted inhibition in insulin, estrogen, beta-estradiol, EGF, EGFR, IL-5, and IL-10RA in the >28-days MV group **(Supplementary Table 9)**. These differences suggest that, at the stage that we sampled the lower airways of patients with critically COVID-19, an overt inflammatory tone was not predictive of worst outcome.

**Figure 5.**
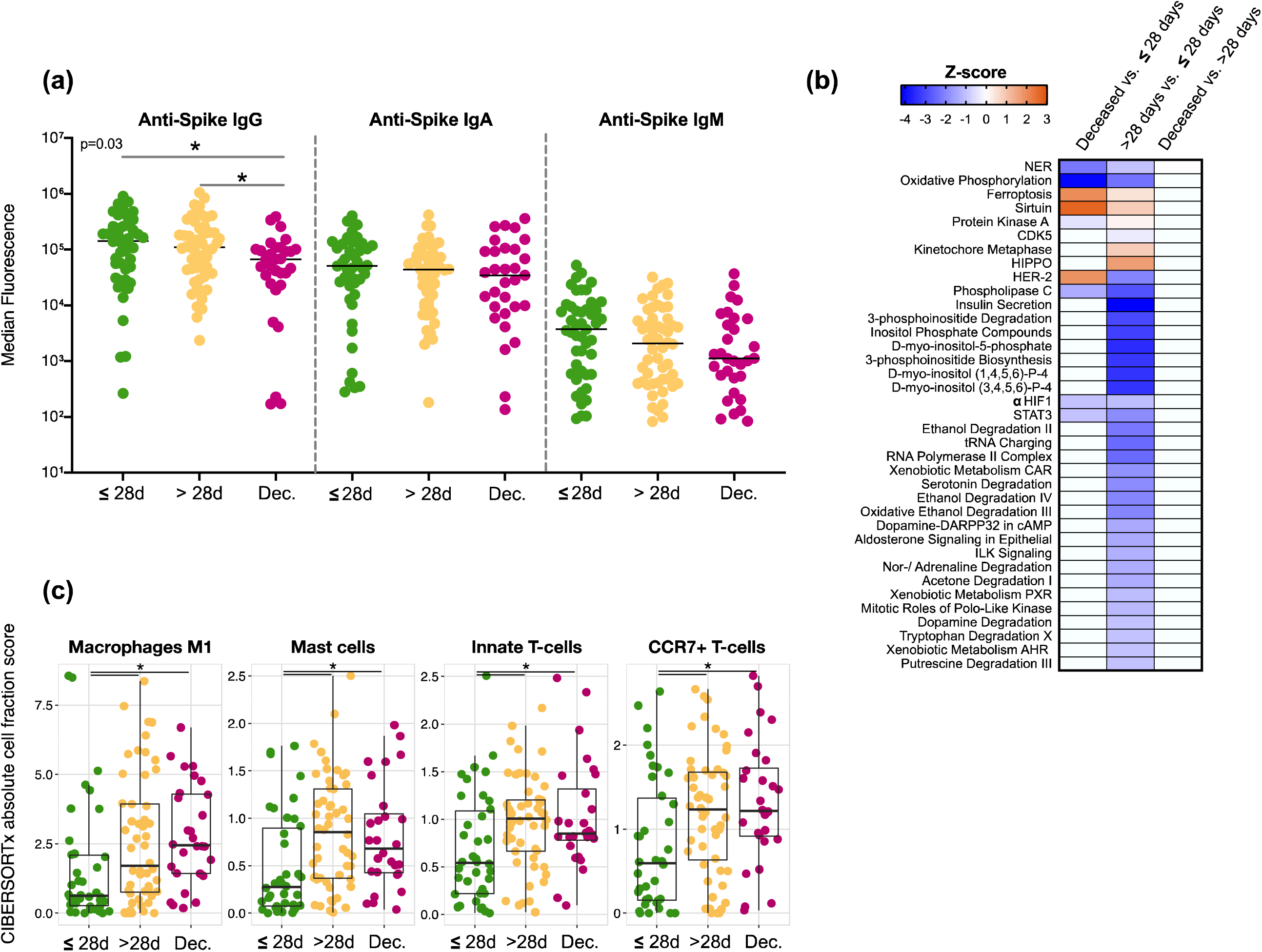
Lower airway host immune profiling in severely ill COVID-19. **(a)** Levels of anti-SARS-CoV-2 Spike antibodies in BAL (*= Mann–Whitney U p<0.05)**. (b)** Heat-map of canonical pathway analysis based on Ingenuity Pathway Analysis (IPA, RRID:SCR_008653) using the lower airway host transcriptome comparing clinical outcome groups. Orange shows up-regulation of pathway, blue shows down-regulation of pathway**. (c)** Cell type abundance quantification plots. Comparison of abundance of mast cells and neutrophils among outcome groups in the BAL fluids of critically ill patients with COVID-19. Cell type abundance was estimated from the host transcriptome with CIBERSORTx. Each dot denotes the quantification score of a sample and boxes depict median and inter-quartile range (*= Mann–Whitney U p<0.05).

To determine if the abundance of immune cells varies between different clinical outcome groups, we estimated cell type abundance from the host transcriptome with computational cell type quantification methods, including a deconvolution approach implemented in CIBERSORTx ^29^ and a cell type signature enrichment approach implemented in xCell ^30^. As reported recently in other studies^31^, among the cell types detected in the BAL samples we observed a consistent enrichment of mast cells and neutrophils in the >28-days MV and deceased groups compared with the ≤28-days MV group (**Figure 5c** and **Supplementary Table 10**). We also identified significantly higher inflammatory macrophages (M1), innate T-cells and memory T-cells (CCR7^+^) among subjects with worse clinical outcome.

### Cross-kingdom network analyses identify bacteria, fungi, and host pathways functionally impacted by SARS-CoV-2

To identify potential microbe-microbe and microbe-host interactions that could have an effect on outcome, we used a multi-scale network analysis approach (Multiscale Embedded Gene co-Expression Network Analysis, MEGENA)^32^. We first used the relative abundance from the RNAseq data to capture co-expressing taxa in the metatranscriptome network neighborhood of SARS-CoV-2 (SARS2-NWN). We examined five such network neighborhoods (constructed by including nodes with increasing distance 1 to 5 from SARS-CoV-2, i.e. neighborhood 1 to neighborhood 5) that were significantly enriched for taxa functionally active in the deceased group when compared with the ≤28-day MV group. Only the largest cluster, with 504 taxa, had significantly enriched taxa in both the deceased and in the ≤28-day MV outcome groups (**Supplementary Fig. 12a**) (FET P-value = 4.6e-45, 4.0 FE). Many of these taxa are among the top 50 most abundant microbes we had previously identified in the metatranscriptome dataset. Taxa present that are influenced by SARS-CoV-2 and significantly differentially enriched in the deceased group include bacteria such as *M. salivarium, Bifidobacterium breve,* and *Lactobacillus rhamnosus* (a gut commensal), that we had previously identified by differential expression analysis (**Figure 3e**), but also taxa such as *S. epidermis*, *Mycoplasma hominis* (urogenital bacteria), and the phage VB_PmiS-Isfahan (also referred to as Proteus virus Isfahan) that we had previously only picked up as being highly abundant but not necessarily differentially enriched in the deceased group. Most of the fungi, such as *C. albicans*, *C. glabrata* and *C. orthopsilosis* were enriched in the ≤28-day MV group. Interestingly, our earlier analysis of the metagenome (**Supplementary Fig. 8b**) had identified *C. glabrata* as being enriched in the deceased group with no enrichment in the metatranscriptome. This analysis indicates that some of these abundant taxa could be responding to SARS-CoV-2 disruption in a similar manner, or indirectly interacting functionally.

We further investigated the association of the network neighborhood with host network modules using the host transcriptome data to identify groups of host genes that are co-expressed in response to SARS-CoV-2 disruption. The 3 host modules with the most significant correlations to SARS2-NWN are M175, M277 and M718. M277 is the parent module of M718, and both are enriched with genes related to respiratory electron transport, while M175 is enriched for IFN-γ signaling (**Supplementary Fig. 12b**). Module M175 is positively correlated with the SARS2-NWN (ρ = 0.32, P-value = 2.1e-3). While there was no collective enrichment of the module by differentially expressed genes (DEGs) in the deceased vs ≤28-days MV, there was for >28-days vs ≤28-days MV (FET P-value = 0.030, 4.5 FE). This module includes well-known antiviral IFN stimulated genes (ISGs), such as *IRF7* and *OASL*. Investigating module response on an individual gene level, Interleukin 4 induced 1 (IL4I1) appears as one of the most up-regulated genes in this module when comparing the deceased group with the ≤28-day MV group. The transporter 1, ATP binding cassette subfamily B member (TAP1) is also upregulated and a key regulator (hub gene). Together with *TAP2*, *TAP1* plays a central role in MHC I antigen presentation^33^. Transcriptional regulators *SP110* and *SP140*, both ISGs and also identified as hub genes, were down-regulated. Module 718 was also positively correlated with the SARS2-NWN (ρ = 0.31, P-value = 1.3e-3; enrichment FET P-value = 0.029, 3.7 FE of M178 by differentially expressed genes in deceased vs ≤28-days MV). The majority of genes in this module are down-regulated in the deceased group compared with the ≤28-day group. Some of the genes encode subunits of the mitochondrial ATP synthase, such as *ATP6* and *ATP8*, the cytochrome C oxidase, with *COX2* and *COX3* as well as the NADH dehydrogenase complex, such as *ND1-ND6*. *ND4L, ATP6, COX2, ND1, ND3, ND4L* and *ND6* are key regulators, potentially modulating the expression of the other genes in the module. These findings further support mitochondria dysfunction^26^, potentially disrupting processes indicated by the module. Other down-regulated genes are humanin1 (*MTRNR2L1*) and R-spondin 1 (*RSPO1*). Humanin is known to protect against oxidative stress and mitochondrial dysfunction^34^. *RSPO1* protects against cell stress by activating the Wnt/β-catenin signaling pathway^35^. Non-coding RNAs, such as *MALAT1* and *RHOQ-AS*1 were found to be up-regulated. *MALAT1* is known to suppress IRF3-initiated antiviral innate immunity^36^ while the function of *RHOQ-AS1* is unknown.

### Metatranscriptome and Transcriptome signatures are predictive of mortality

We evaluated the strength of the metatranscriptomic, metagenomic and host transcriptomic profiles to predict mortality in this cohort of critically ill COVID-19 patients. To this end, we identified features in each of these datasets and constructed risk scores that best predicted mortality. **Figure 6a** shows that the metatranscriptome data, alone or combined with the other two datasets, was most predictive of mortality. Importantly, the predictive power (as estimated by the area under the curve) of the metatranscriptome data was improved by excluding probable contaminants and worsened when SARS-CoV-2 was removed from the modeling. The selected features we used to construct the metatranscriptome, metagenome and host transcriptome risk scores are reported in **Supplementary Table 11**). Using the means of the scores, we classified all subjects into high risk and low risk groups for mortality. **Figure 6b** shows Kaplan-Meier survival curve comparisons evaluating the predictive power of risk score stratification based on metatranscriptome, metagenome and host transcriptome data. Combining risk scores from different datasets showed an optimal identification of mortality when metatranscriptome and host transcriptome were considered (**Figure 6c**). We then used the gene signature found as being the most predictive of mortality to conduct IPA analyses. Among the upstream regulators, mortality was associated with predicted activation of interferon alpha while chemotaxis and infection by RNA virus were predicted as activated in diseases and functions. These data highlight the importance of SARS-CoV-2 abundance in the lower airways as a predictor for mortality, and the significant contribution of the host cell transcriptome, which reflects the lower airway cell response to infection.

**Figure 6.**
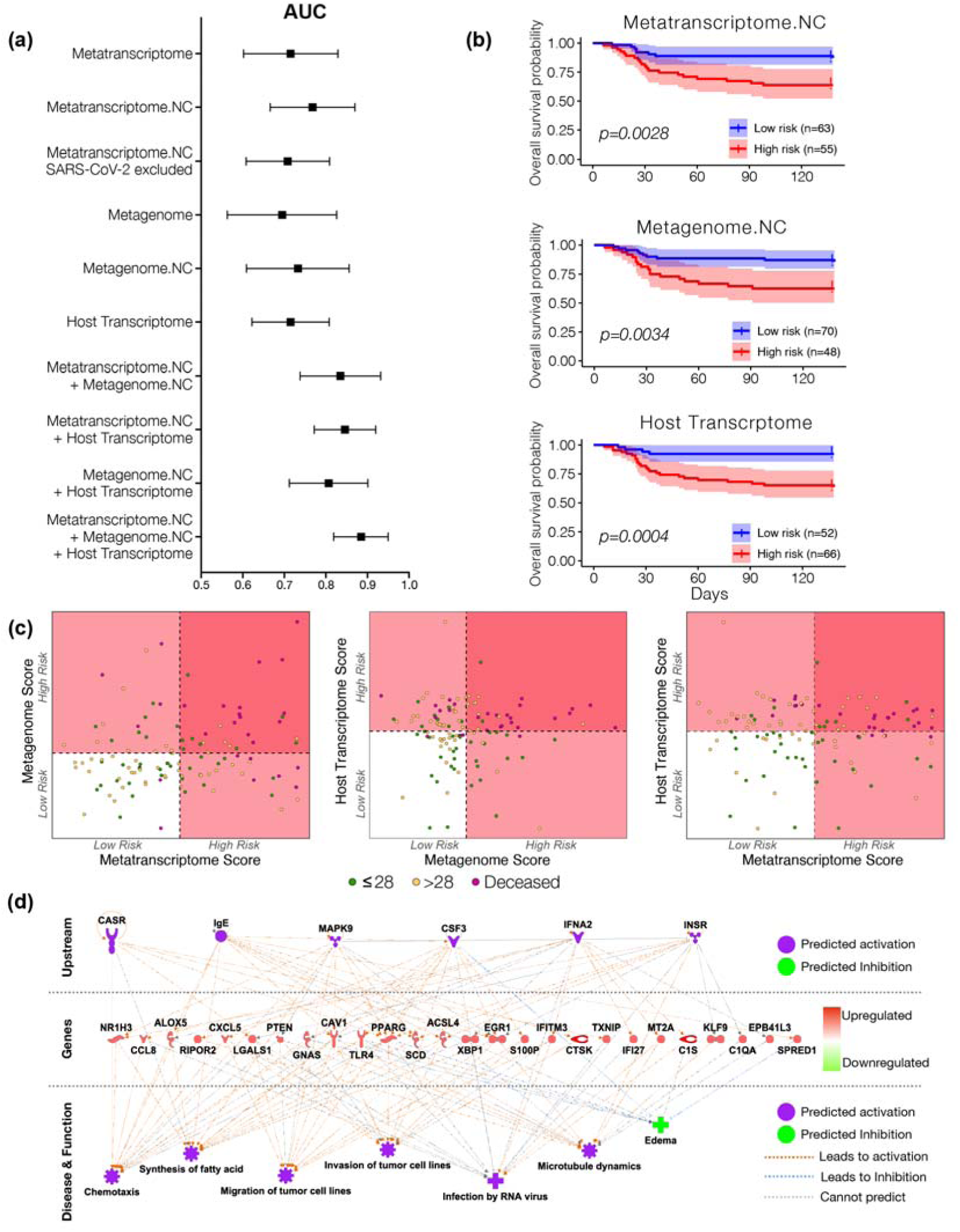
Mortality predictive power of metatranscriptome, metagenome and host transcriptome. **(a)** Area under the curved median and confidence interval for receiver operating characteristic curve analyses calculated from each sequencing datasets as predictor and mortality as outcome. **(b)** Kaplan-meier survival analyses based on a cutoff value estimated from features selected from each sequencing dataset. The “High risk” and “Low risk” groups is the mean of predicted risk scores in all samples. **(c)** Scatterplot among risk scores from metatranscriptome, metagenome, and host transcriptome. Dotted line denotes the mean of the risk scores across all subjects, which is also the threshold for dividing the samples into “High risk” and “Low risk” groups. **(d)** IPA analyses of host transcriptomic signatures identified as most predictive of mortality.

## Discussion

A limited number of studies to date have evaluated the lower airway microenvironment in patients with SARS-CoV-2 infection because of the increased risk of virus transmission to healthcare providers during sampling^19,20, 37–42^. This has limited molecular investigations into the primary site of the disease. Having built a substantial biorepository of lower airway samples among COVID-19 patients on mechanical ventilation recruited during the first wave of SARS-CoV-2 infections in New York City, we used a metagenomic approach to characterize the microbiome in the lower airways and assessed its impact on clinically meaningful outcomes. In this analysis of 142 critically ill hospitalized patients with confirmed SARS-CoV-2 infection and lower airway biorepository samples available, we determined that higher SARS-CoV-2 viral load, higher relative abundance of *Mycoplasma salivarium*, and limited anti-SARS-CoV-2 Spike protein IgG response in the lower airways were associated with increased mortality. This signature was supported by the metatranscriptome data of the lower airway samples where SARS-CoV-2 sequence reads were significantly enriched in those patients who died compared to those who survived after developing respiratory failure requiring mechanical ventilation. Importantly, although we observed changes in other microbial components of the lower airway microbiome in our analysis of lower airway samples from 118 patients and by clinical laboratory culture results obtained from 589 patients, we did not find evidence to support the hypothesis that co-infection with common (bacterial, viral, fungi) respiratory pathogens was associated with poor outcome—although most patients received empiric treatment with broad spectrum antibiotics and anti-fungals.

Several studies have explored the relationship between SARS-CoV-2 viral load and mortality^43–48^. Severe influenza requiring hospitalization has also been associated with higher viral loads^49,50^. It has been argued that high viral load might merely be a reflection of an individual’s immune response^43^. In fact, in SARS-CoV-1, clinical progression was not associated with increased viral load or uncontrolled viral replication in the nasopharynx but rather with an upregulated immune profile in these patients^51^. In a large cohort of 1145 patients with confirmed SARS-CoV-2, viral load measured in nasopharyngeal swab samples was found to be significantly associated with mortality, even after adjusting for age, sex, race and several co-morbidities ^48^. Similar results were found in a cohort of patients in New York City with or without cancer, where in-hospital mortality was significantly associated with a high SARS-CoV-2 viral load in the upper airways ^47^. The data presented here through the use of direct quantitative methods (RT-PCR) and a semiquantitative untargeted approach (metatranscriptome sequencing) support the hypothesis that the SARS-CoV-2 viral load in the lower airways plays a critical role in the clinical progression of critically ill COVID-19 patients. It is important to note that current guidelines for treatment of COVID-19 do not recommend treatment with remdesivir for patients receiving invasive mechanical ventilation^52^. The results of this investigation suggest that antivirals might still have a role in the treatment of critically ill COVID-19 patients.

We investigated the possibility that mortality with SARS-CoV-2 infection was related to co-infection with other pathogens. To this point several investigations have shown evidence of SARS-CoV-2 co-infection with other viruses, bacteria and fungi identified by culture-based techniques ^18,53–60^. In a cohort of 116 specimens positive for SARS-CoV-2, 21% were positive for one or more additional respiratory pathogens including rhinovirus/enterovirus and respiratory syncytial virus^53^. In a meta-analysis of 3,338 patients with COVID-19, only 3.5% of patients had an identified bacterial co-infection at admission, while 14.3% were found later to have a secondary bacterial infection^55^. The most common pathogens identified included species in the genera *Mycoplasma*, *Hemophilus,* and *Pseudomonas*. In another study, the most commonly identified co-infections were with *Streptococcus pneumoniae*, *Klebsiella pneumoniae*, and *Haemophilus influenzae*^57^. Using detailed clinical laboratory culture data available for 589 subjects hospitalized with respiratory failure due to COVID-19, we showed that higher rates of respiratory infection with other organisms, especially early in their hospitalization, did not occur among subjects with poor clinical outcome. Further, we did not observe an association between positive cultures for any pathogen tested and increased odds of dying in critically ill COVID-19 patients.

In the subset of COVID-19 patients with BAL samples, we used NGS to identify all potential pathogens and commensals in the lower airways beyond microbial cultures routinely obtained as per clinical care. The RNA virome data showed that SARS-CoV-2 dominates the lower airways and was significantly associated with death. A small number of samples had a few sequences that mapped to influenza A or B viruses, suggesting that co-infection with influenza did not occur frequently during this first wave of SARS-CoV-2 infections. Within the DNA virome, there was no significant difference in viruses between the three outcome groups despite the frequent finding of HSV-1. Similarly, when evaluating the metatranscriptome of DNA viruses, there were few differences between the three outcome groups. Although analysis of the phage metagenome data showed no differential enrichment between the three cohorts, we did identify in the metatranscriptome data differentially active phages when comparing the three cohorts, suggesting that changes in the bacterial microbiome may be occurring in critically ill patients with COVID-19. Certain *Staphylococcus* phages were differentially active in those who were ventilated for more than 28 days and in those who died. Interestingly, the bacterial signatures also identified *Mycoplasma salivarium*, a known oral commensal that has previously been associated with ventilator-acquired pneumonia^61^, as differentially active in those who died and those who were ventilated for more than 28 days when compared to those ventilated less than 28 days. From previous data published by us, enrichment of the lower airway microbiota with oral commensals was seen to be associated with a pro-inflammatory state in several diseases including lung cancer^62,63^ and non-tuberculosis mycobacterium related bronchiectasis^64^.

With the use of metagenomic and metatranscriptomic analyses it is also possible to examine how functionally active microbes impact the host^65^. In this cohort of patients, we evaluated the functional profile of the microbiome within the lower airways and its effect on mortality, something that, to our knowledge, had not yet been assessed in COVID-19 patients. The only significant gene function enrichment was found with the metatranscriptome data suggesting that functional activation of microbes can provide further insights into the lower airway microbial environment of patients with worst outcome. Among the pathways that were differentially expressed in those patients with poor outcome, we identified genes associated with degradation, transport, and antimicrobial resistance genes, as well as with signaling. These differences may indicate important functional differences leading to a different metabolic environment in the lower airways that could impact host immune responses. It could also be representative of differences in microbial pressure in patients with higher viral loads and different inflammatory environments.

In the current investigation, we also characterized the immune response within the lower airways by measuring anti-SARS-CoV-2 Spike antibodies and profiling the host RNA transcriptome. We observed that low levels of anti-Spike and anti-RBD IgG in the lung were associated with poor outcome. Although we did not find a statistically significant association between SARS-CoV-2 neutralizing capacity and poor outcome, levels of SARS-CoV-2 neutralizing antibodies, anti-Spike and anti-RBD antibodies (both IgG and IgA) were negatively correlated with SARS-CoV-2 viability. Prior investigations have suggested that IgA levels are a key driver of neutralization in the mucosa^66–68^. The differences noted in the current investigation in the IgG pools are intriguing and future work investigating the antibodies generated during SARS-CoV-2 infections will be essential.

When examining host transcriptomic differences across the different clinical outcome groups, Sirtuin and Ferroptosis signaling pathways were found to be upregulated in the most critically ill COVID-19 patients. Upregulation in the Sirtuin pathway demonstrates an increased host inflammatory response to viral infection^23^. In addition, ferroptosis, a recently identified form of non-apoptotic regulated cell death through iron-dependent accumulation of lipid peroxides, has been shown to cause direct lung injury^69^ or pulmonary ischemia-reperfusion injury^70,71^. Interestingly, there is evidence to support that STAT3^69^ and ACSL4^70^ alleviated ferroptosis-mediated acute lung injury dysregulation, which are both down-regulated in COVID-19 patients with worse clinical outcome. Further analysis showed that there appeared to be an inactivation of phagocytes, neutrophils, granulocytes, and leukocytes, including downregulation of IgG expression levels, with additional mitochondria dysfunction, and down-regulation of Inositol related pathways and noradrenaline/adrenaline degradation. There is evidence that in the neonatal lung, inositol related components exert an anti-inflammatory effect and can prevent acute lung injury^72,73^.

Collectively, these data suggest that an imbalance rather than an elevated inflammatory state in the lung is an important marker that predicts poor outcomes in critically ill COVID-19 patients. Indeed, the inferred cell composition analysis from the bulk transcriptome data overall points to a tepid immune response. Memory T cells have been implicated with a robust immune response in SARS-CoV-2.^74^ The deficiency of these memory T cells that we found in the lungs of COVID-19 patients with worse outcome further supports the presence of an ineffective immune response or presence of immune exhaustion. IL4I1, found in the network analysis to be up-regulated in the deceased group in association with SARS-CoV-2, is an immunosuppression enzyme that plays a role in infection and the control of immunopathology^75^. IL4I1 induction has been reported in viral infections with influenza virus^76^. The ISGs and transcriptional regulators *SP110* and *SP140*, both downregulated in the deceased group. play important roles in resisting intracellular pathogens^77^.

Strikingly, interrogation of the host transcriptomic analysis identified survival-associated differences in interferon-related responses. Our host transcriptomic risk stratified model seems to point to a predictive activation of type I interferon as a prediction for mortality. This might be inconsistent with the current suggestion that, based on systemic levels, early interferon responses are associated with poor outcome in COVID19.^78,79^ Others have suggested that a robust interferon response may lead to a hyperinflammatory state that could be detrimental in the disease process, justifying the use of Janus kinase inhibitor inhibitors in patients with COVID-19.^80^ Studies comparing transcriptomic signatures in BAL of patients with severe COVID-19 and controls have shown activation of type 1 interferons.^81^ While further longitudinal data will be needed to clarify the role of interferon signaling on the disease, the data presented here suggest that combining microbial and host signatures could help understand the increase risk for mortality in critically ill COVID-19 patients.

By collecting BAL samples rather than endotracheal aspirate specimens we were able to ensure extensive sampling of the lower respiratory tract in intubated patients. However, we were limited to samples from intubated patients in whom a clinically indicated bronchoscopy was done to place a percutaneous tracheostomy or for airway clearance. Although this included a large number of patients with various clinical outcomes, those sampled may not be representative of the extremes in the spectrum of disease severity who were most likely not eligible for bronchoscopy. For example, patients that presented with very rapid clinical deterioration and died within the first few days of hospitalization or those who were quickly weaned from mechanical ventilation did not receive bronchoscopy. However, extensive and detailed clinical data were also obtained from intubated COVID-19 patients without bronchoscopy performed within the Manhattan Campus (no bronchoscopy cohort) and from the Long Island cohort for whom bronchoscopies were done without collecting research samples. In both of these cohorts, clinical laboratory culture data did not identify untreated secondary pathogen infections associated with poor outcome.

The samples used in this investigation were obtained during the first surge of cases of COVID-19 in New York City, and management reflected clinical practices at that time. Among the differences with current therapeutic approaches in COVID-19 patients, corticosteroids and remdesivir, two medications that likely affect the lower airway microbial landscape, were rarely used during the first surge. Other medications, such as antibiotics and anti-inflammatory drugs could affect our findings and we therefore considered them as potential confounders. However, the use of these medications was not found to be associated with clinical outcome. The cross-sectional study design precluded evaluation of the temporal dynamics of the microbial community or the host immune response in this cohort, which could provide important insights into the pathogenesis of this disease. Performing repeated bronchoscopies without a clinical indication would be challenging in these patients and other less invasive methods might need to be considered to study the lower airways at earlier timepoints and serially over time in patients with respiratory failure. It is important to note that there were no statistically significant differences in the timing of sample collection across the three outcome groups.

In summary, we present here the first evaluation of the lower airway microbiome using a metagenomic and metatranscriptomic approach, along with host immune profiling in critically ill patients with COVID-19 requiring invasive mechanical ventilation. The RNA metatranscriptome analysis showed an association between the abundance of SARS-CoV-2 and mortality, consistent with the signal found when viral load was assessed by targeted rRT-PCR. These viral signatures correlated with lower anti-SARS-CoV-2 Spike IgG and host transcriptomic signatures in the lower airways associated with poor outcome. Importantly, both through culture and NGS data, we did not find evidence for an association between untreated infections with secondary respiratory pathogens and mortality. Together, these data suggest that active lower airway SARS-CoV-2 replication and poor SARS-CoV-2-specific antibody responses are the main drivers of increased mortality in COVID-19 patients requiring mechanical ventilation. The potential role of oral commensals such as *Mycoplasma salivarium* need to be explored further. It is possible that *M. salivarium* can impact key immune cells and has recently been reported at a high prevalence in patients with ventilator-acquired pneumonia^61^. Critically, our finding that SARS-CoV-2 evades and/or derails effective innate/adaptive immune responses indicates that therapies aiming to control viral replication or induce a targeted antiviral immune response may be the most promising approach for hospitalized patients with SARS-CoV-2 infection requiring invasive mechanical ventilation.

## Methods

### Subjects

Enrolled subjects were 18 years or older, admitted to the intensive care units (ICUs) at NYU Langone Health from March 10^th^ to May 10^th^, 2020 with a nasal swab confirmed diagnosis of SARS-CoV-2 infection by reverse transcriptase polymerase chain reaction (RT-PCR) assay and respiratory failure requiring invasive mechanical ventilation. Samples were obtained during clinically indicated bronchoscopy performed for airway clearance or for percutaneous tracheostomy placement. Surviving subjects signed informed consent to participate in this study. Samples and metadata from subjects who died or were incapacitated were de-identified and included in this study. Comprehensive demographic and clinical data were collected. We also collected longitudinal data on clinical laboratory culture results and treatment. **Supplementary figure 1** shows the distribution of subjects and sampling strategy used for this study. The study protocol was approved by the Institutional Review Board of New York University.

### Lower airway bronchoscopic sampling procedure

Both background and supraglottic (buccal) samples were obtained prior to the procedure, as previously described^62^. The background samples were obtained by passing sterile saline through the suctioning channel of the bronchoscope prior to the procedure. Bronchoalveolar lavage (BAL) samples were obtained from one lung segment as per discretion of the treating physician as clinically indicated. Samples were then transferred to a BSL3 laboratory for processing. Once there, 2 mL of whole BAL was stored in a tube prefilled with 2 mL of Zymo Research’s DNA/RNA Shield™ (R1100-250, https://www.zymoresearch.com/pages/covid-19-efforts) for RNA/DNA preservation and virus inactivation. In addition, background control samples (saline passed through the bronchoscope prior to bronchoscopy) and supraglottic aspirates were stored in the same RNA/DNA shield. A subset of samples underwent BAL cell separation by centrifugation and cells were cryopreserved in DMSO while acellular BAL fluid was aliquoted for cytokine measurements. A paired blood sample was also obtained in EDTA tubes (Becton Dickinson, ref# 366450) and PAXgene Blood RNA tubes (PreAnalytiX) ref# 762165).

### Viral load detection targeting the N gene

SARS-CoV-2 viral load was measured by quantitative real-time reverse transcription polymerase chain reaction (rRT-PCR) targeting the SARS-CoV-2 nucleocapsid (N) gene and an additional primer/probe set to detect the human RNase P gene (RP). Assays were performed using Thermo Fisher Scientific (Waltham, MA) TaqPath 1-Step RT-qPCR Master Mix, CG (catalog number A15299) on the Applied Biosystems (Foster City, CA) 7500 Fast Dx RealTime PCR Instrument. Using the positive controls provided by the CDC, which are normalized to 1000 copies/mL, we converted the different Ct positive to copies/mL. This was done using the DDCT method, applying the formula: Power [2, (CT (sample, N1 gene) - CT (PC, N1 gene)] – [CT (sample, RP gene) - CT (PC, RP gene)]*1000.

### SARS-CoV-2 viral viability through measurement of subgenomic transcripts

Viral subgenomic mRNA (sgRNA) is transcribed in infected cells and is not packaged into virions. Thus, presence of sgRNA is indicative of active infection of a mammalian cell in samples. We therefore measure sgRNA in all BAL samples obtained targeting the E gene as previously described.^21^ Briefly, five µl RNA was used in a one-step real-time RT-PCR assay to sgRNA (forward primer 5’-CGATCTCTTGTAGATCTGTTCTC-3’; reverse primer 5’-ATATTGCAGCAGTACGCACACA-3’; probe 5’-FAM-ACACTAGCCATCCTTACTGCGCTTCG-ZEN-IBHQ-3’) and using the Quantifast Probe RT-PCR kit (Qiagen) according to instructions of the manufacturer. In each run, standard dilutions of counted RNA standards were run in parallel to calculate copy numbers in the samples.

### DNA/RNA isolation, library preparation and sequencing

DNA and RNA were isolated in parallel using zymoBIOMICS™ DNA/RNA Miniprep Kit (Cat: R2002) as per manufacturer’s instructions. DNA was then used for whole genome shotgun (WGS) sequencing using it as input into the NexteraXT library preparation kit following the manufacturer’s protocol. Libraries were purified using the Agencourt AMPure XP beads (Beckman Coulter, Inc.) to remove fragments below 200 bp. The purified libraries were quantified using the Qubit dsDNA High Sensitivity Assay kit (Invitrogen) and the average fragment length for each library was determined using a High Sensitivity D1000 ScreenTape Assay (Agilent). Samples were added in an equimolar manner to form two sequencing pools. The sequencing pools were quantified using the KAPA Library Quantification Kit for Illumina platforms. The pools were then sequenced on the Illumina Novaseq 6000 in one single run. For RNA sequencing, RNA quantity and integrity were tested with a BioAnalyzer 2100 (Agilent). Among bronchoscope control (BKG) samples, only 5 yielded RNA with sufficient quality and quantity to undergo library preparation and sequencing. The automated Nugen Ovation Trio Low Input RNA method was used for library prep with 3ng total RNA input of each sample. After 6 amplification cycles, samples were sequenced using 2x Novaseq 6000 S4 200 cycle Flowcells using PE100 sequencing.

### Microbial community characterization using whole genome shotgun sequencing (WGS) and RNA metatranscriptome

For all metagenomic and metatranscriptomic reads, Trimmomatic v0.36^82^, with leading and trailing values set to 3 and minimum length set to 36, was used to remove adaptor sequences. All rRNA reads were then removed from the metatranscriptomic reads using SortMeRNA v4.2.0^83^ with default settings. Metagenomic and filtered metatranscriptomic reads were mapped to the human genome using Bowtie2 v2.3.4.1^84^ with default settings and all mapping reads were excluded from subsequent microbiome, mycobiome, and virome metagenomic and metatranscriptomic analysis. Technical replicates for each biological sample were pooled together for subsequent analyses. Taxonomic profiles for all metagenomic and metatranscriptomic samples were generated using Kraken v2.0.7^85^ and Bracken v2.5 [https://doi.org/10.7717/peerj-cs.104] run with default settings. The database used for quantifying taxonomic profiles was generated using a combined database containing human, bacterial, fungal, archaeal, and viral genomes downloaded from NCBI RefSeq on January 8, 2021. Additionally, genomes for Candida auris (Genbank: GCA_003013715.2, GCA_008275145.1) and Pneumocystic jirovecii (Genbank: GCA_001477535.1) were manually added to the database. Differentially abundant bacterial and viral taxa were identified for the BAL and UA samples groups individually using DESeq2 v1.28.1^86^ with the three group clinical outcome meta-data readouts set as the sample groupings. Significantly differentially abundant taxa contained at a minimum an aggregate of 5 reads across samples and had an FDR <0.2^87,88^.

For functional microbial profiling, processed sequencing reads were further depleted of human-mapping reads by removing all reads classified as human by Kraken v2.0.7^85^ using KrakenTools v0.1-alpha (https://github.com/jenniferlu717/KrakenTools). FMAP v0.15^89^ was run on both the metagenomic and metatranscriptomic reads to profile the metabolic pathways present in each sample. FMAP_mapping.pl paired with diamond v0.9.24^90^ and FMAP_quantification.pl were used with default settings to identify and quantify proteins in the Uniref90 database. Using DESeq2 v1.28.1^86^, differentially expressed genes were identified for the BAL samples individually using the three group clinical outcome-metadata readouts for all genes that had an aggregate 5 reads across all samples.

Antibiotic resistance genes were quantified in all metagenome and metatranscriptome samples using Salmon v1.3.0^91^ run with --keepDuplicates for indexing and --libtype A -- allowDovetail --meta for quantification. Genes were filtered such that only genes that actively conferred antibiotic resistance were kept. To assess differentially expressed classes of antibiotic resistance genes, gene counts for individual antibiotic resistance genes were collapsed by their conferred antibiotic resistance.

**Supplementary Figure 1** shows a summary of depth achieved with the parallel WGS and metatranscriptome approach across sample types and the number of reads assigned to different microbial subfractions (bacteria, fungi, DNA viruses, RNA viruses and phages). Further analysis was also done to identify possible contaminants in the metatranscriptome and metagenome datasets. To this end, we compared the relative abundance of taxa between background bronchoscope control and BAL samples. Taxa with median relative abundance greater in background than in BAL were identified as probably contaminant and listed in **Supplementary Table 4).** None of the taxa identified as possible contaminants were removed from the analyzed data but are shown for comparison with signatures identified in the rest of the analyses.

### Anti-Spike SARS-CoV-2 antibody profiling in BAL

BAL samples were heat-treated at 56°C for one hour, and centrifuged at 14000g for 5 min. The supernatant was collected and diluted 50-fold in PBST containing 1% skim milk. The diluted samples were incubated at room temperature (R.T.) for 30 min with QBeads DevScreen: SAv (Streptavidin) (Sartorius 90792) that had been loaded with biotinylated Spike, biotinylated RBD or biotin (negative control) in wells of a 96 well HTS filter plate (MSHVN4550). As positive controls, we used CR3022 antibody, that recognizes SARS-CoV-2 Spike and RBD, in human IgG, IgA and IgM formats (Absolute Antibody). After washing the beads, bound antibodies were labeled with anti IgG-DyLight488, anti IgA-PE and anti IgM-PECy7, and the fluorescence intensities were measured in Intellicyt IQue3 (Sartorius). The acquired data [median fluorescence intensity (MFI)] were normalized using the MFI values of the CR3022 antibodies to compensate for variations across plates. **Supplementary Figure 10** shows that the levels of these antibodies were higher in BAL samples of patients with SARS-CoV-2 than in BAL samples from 10 uninfected healthy smokers recruited for research bronchoscopy. Details of method development and validation will be described elsewhere (Koide et al. in preparation).

### SARS-CoV-2 preparation and neutralization assay

icSARS-CoV-2-mNG (isolate USA/WA/1/2020, obtained from the UTMB World Reference Center for Emerging Viruses and Arboviruses) was amplified once in Vero E6 cells (P1 from the original stock). Briefly, 90-95% confluent T175 flask (Thomas Scientific) of Vero E6 (1×10^7^ cells) was inoculated with 50 μL of icSARS-CoV-2-mNG in 5 mL of infection media (DMEM, 2% FBS, 1% NEAA, and 10 mM HEPES) for 1 hour. After 1 hour, 20 mL of infection media was added to the inoculum and cells were incubated 72 hours at 37 °C and 5% CO_2_. After 72 hours, the supernatant was collected and the monolayer was frozen and thawed once. Both supernatant and cellular fractions were combined, centrifuged for 5 min at 1200 rpm, and filtered using a 0.22 μm Steriflip (Millipore). Viral titers were determined by plaque assay in Vero E6 cells. In brief, 220,000 Vero E6 cells/well were seeded in a 24 well plate, 24 hours before inoculation. Ten-fold dilutions of the virus in DMEM (Corning) were added to the Vero E6 monolayers for 1 hour at 37 °C. Following incubation, cells were overlaid with 0.8% agarose in DMEM containing 2% FBS (Atlanta biologicals) and incubated at 37 °C for 72 h. The cells were fixed with 10% formalin, the agarose plug removed, and plaques visualized by crystal violet staining. All procedures including icSARS-CoV-2-mNG virus were performed using Biosafety Level 3 laboratory conditions.

For SARS-CoV-2 neutralization assays, Vero E6 cells (30,000 cells/well) were seeded in a 96 well plate 24 h before infection. Two-fold serial dilutions of BAL lysates were mixed with mixed 1:1 (vol/vol) with SARS-CoV-2 mNG virus (multiplicity of infection, MOI 0.5), and incubated for 1 h at 37 °C. After incubation, 100 μL of the mixtures of the antibody and SARS-CoV-2 mNG were added to the Vero E6 monolayers, and cells were incubated at 37°C. After 20 h, cells were fixed with 4 % formaldehyde (Electron Microscopy Sciences) at room temperature for 1 h. After fixation, cells were washed twice with PBS and permeabilized with 0.25% triton-100, stained with DAPI (Thermo), and quantified on a CellInsight CX7 High-content microscope (Thermo) using a cut-off for three standard deviations from negative to be scored as an infected cell.

### Transcriptome of BAL cells

RNA-Seq was performed on bronchial epithelial cells obtained by airway brushing, as described^92–94^, using the Hi-seq/Illumina platform at the NYU Langone Genomic Technology Center (data available at Sequence Read Archive: # PRJNA592149). KEGG^95,96^ annotation was summarized at levels 1 to 3. Genes with an FDR-corrected adjusted p-value <0.25 were considered significantly differentiated, unless otherwise specified. Pathway analysis using differentially regulated genes (FDR<0.25) was done using Ingenuity Pathway Analysis, RRID:SCR_0-at least 1 count per million in at least two samples were retained. For digital cytometry with CIBERSORTx, a signature matrix derived from single-cell transcriptome of BAL cells collected from patients with COVID-19^31^ was first generated with the “Create Signature Matrix” module in the CIBERSORTx online tool. A maximum of 10 cells per cell type per patient were initially sampled from the original data and 20 cells per cell type were then used to build the single-cell reference with the default parameters. Then the “Impute Cell Fractions” module was used to estimate the absolute cell fraction score of different cell types in bulk transcriptomes using the single-cell signatures with “S-mode” batch correction and 100 permutations in the absolute mode. Bulk transcriptomes with a significant deconvolution p-value (≤0.05) were retained. For xCell cell type signature enrichment analysis, the enrichment scores were inferred with built-in signature of cell types detected in the BAL samples as reported previously ^31^. The two-tailed Wilcoxon rank sum test with Benjamini-Hochberg correction were computed between groups of samples for comparison.

### Microbial and Host predictive modeling

Cox proportional hazards model was used for investigating the association between the time to death and the relative abundance of each taxon quantified using metatranscriptomic and metagenomic data separately. We first performed the univariate screening test to identify significant features associated with the time to death using the Cox proportion hazards regression model for the relative abundance of taxa from the RNA and DNA data, and log-transformed count of host transcriptome data, respectively. Within each type of data, given the p-value cutoff, the features with a p-value less than the cutoff were selected and integrated as a sub-community. For the RNA and DNA data, the alpha diversity (Shannon index) was calculated for each sample on the selected sub-community and the negative of the value was defined as the microbial risk score, because high alpha diversity indicates low risk of death. For the host transcriptome data, the log-transformed total count of all selected candidate transcriptome for each sample was defined as the risk score, since most selected candidate transcriptomes increased the risk of death. The leave-one-out cross-validation (LOOCV) was used for the predictions. The p value cutoff was set at the value which produces the largest AUC (area under the receiver operating characteristic curve) in predicting the death/survival status using the risk score we constructed over these features. The additive model was used to integrate when more than one scores are used for the prediction.

### Multiscale and co-expression network analyses

Raw counts from the human transcriptome were normalized and converted to log2-counts per million using limma^97^/voom^98^ (v3.44.1 with R v4.0.0) with standard parameters. Microbiome abundance information was converted to relative abundance. Low abundance taxa were removed based on average abundance across all samples to yield a minimum of 1000 taxa for each metatranscriptome dataset. All datasets were batch adjusted. Differentially expressed genes (DEGs) and differentially abundant taxa were called using the DESeq2 package^86^ (v1.28.1), based on the negative binomial (i.e. Gamma-Poisson) distribution. According to the recommendation by the authors, we used non-normalized data (i.e. raw gene counts and abundance data), as DESeq2 internally corrects data and performs normalization steps. For this purpose, raw microbiome abundance data were converted to DESeq2 dds objects using the phyloseq R library (V1.32.0). Contrasts are based on outcome groups (≤28 days MV, > 28 days MV or death). Differentially expressed genes and differentially abundant tax with FDR of 0.2 or less are considered significant.

Multiscale Embedded Gene Co-Expression Network Analysis (MEGENA) ^32^ was performed to identify host modules of highly co-expressed genes in SARS-CoV-2 infection. The MEGENA workflow comprises four major steps: 1) Fast Planar Filtered Network construction (FPFNC), 2) Multiscale Clustering Analysis (MCA), 3) Multiscale Hub Analysis (MHA), 4) and Cluster-Trait Association Analysis (CTA). The total relevance of each module to SARS-CoV-2 infection was calculated by using the Product of Rank method with the combined enrichment of the differentially expressed gene (DEG) signatures as implemented: *G_J_* = Π*_i_g_ji_*, where, *g_ji_* is the relevance of a consensus ***j*** to a signature ***i;*** and *g_ji_* is defined as (*max_j_*(*r_ji_*) + 1 – *r_ji_*)/Σ*_j_ r_ji_*, where *r_ji_* is the ranking order of the significance level of the overlap between the module ***j*** and the signature.

To functionally annotate gene signatures and gene modules derived from the host transcriptome data, we performed an enrichment analysis of the established pathways and signatures↓including the gene ontology (GO) categories and MSigDB. The hub genes in each subnetwork were identified using the adopted Fisher’s inverse Chi-square approach in MEGENA; Bonferroni-corrected p-values smaller than 0.05 were set as the threshold to identify significant hubs. The correlation between modules, modules and clinical traits as well as modules and individual taxa were performed using Spearman correlation. Other correlation measures, such as Pearson correlation or the Maximal Information Coefficient (MIC)^99^ proved to be inferior for this task. Categorical trait data was converted to numerical values as suitable.

### Data availability

Sequencing data are available in NCBI’s Sequence Read Archive under project numbers PRJNA688510 and PRJNA687506 (RNA and DNA sequencing, respectively). Codes used for the analyses presented in the current manuscript are available at https://github.com/segalmicrobiomelab/SARS_CoV2.

## Data Availability

Sequencing data are available in NCBI's Sequence Read Archive at the links provided bellow.

https://www.ncbi.nlm.nih.gov/sra/?term=PRJNA688510

https://www.ncbi.nlm.nih.gov/sra/?term=PRJNA687506

## Acknowledgement

We would like to thank the Genome Technology Center (GTC) for expert library preparation and sequencing, and the Applied Bioinformatics Laboratories (ABL) for providing bioinformatics support and helping with the analysis and interpretation of the data. Experimental Pathology Research Laboratory for histopathology services and imaging. GTC and ABL are shared resources partially supported by the Cancer Center Support Grant P30CA016087 at the Laura and Isaac Perlmutter Cancer Center. This work has used computing resources at the NYU School of Medicine High Performance Computing Facility (HPCF). Financial support for the PACT project is possible through funding support provided to the FNIH by: AbbVie Inc., Amgen Inc., Boehringer-Ingelheim Pharma GmbH & Co. KG, Bristol-Myers Squibb, Celgene Corporation, Genentech Inc., Gilead, GlaxoSmithKline plc, Janssen Pharmaceutical Companies of Johnson & Johnson, Novartis Institutes for Biomedical Research, Pfizer Inc., and Sanofi. The findings and conclusions in this report are those of the authors and do not necessarily represent the official position of the Centers for Disease Control and Prevention.

## Supplementary Figure Legends

**Supplementary Figure 1. Description of patient cohort, samples obtained, analyses performed and sequencing depth**.

**Supplementary Figure 2. SARS-CoV-2 viral load in upper airway samples.** Copies of the SARS-CoV-2 N gene per ml, normalized by the Human RNase P gene, in upper airways comparing clinical outcome groups (Mann–Whitney U p-value).

**Supplementary Figure 3. Identification of top taxa found in background samples as compared with BAL and upper airway samples.** Boxplots showing the relative abundance values in log10 relative abundance of taxa ranked ordered based on dominance of Background bronchoscope control samples and compared to abundances in BAL and Upper Airway samples within metatranscriptome **(a)** and metagenome **(b)** data. Red labels indicate taxa where relative abundance is higher in background samples than in BAL and therefore considered possible contaminant.

**Supplementary Figure 4. Topographical analyses of Metatranscriptome data.** Comparison of alpha diversity (Shannon Index, **a**) and beta diversity (Bray Curtis Dissimilarity index, **b**) across background negative controls (bronchoscope), bronchoalveolar lavage (BAL) and upper airway (UA) samples (Kruskal-Wallis and PERMANOVA p-values, respectively). **(c)** Boxplots showing the relative abundance values in log10 across all metatranscriptome samples for the BAL and Upper Airway samples. The 50 taxa with the highest relative abundance values in the BAL metatranscriptome data are displayed; the top 10 in the BAL are highlighted in bold. Each column consists of four plots displaying in order from top to bottom, the most abundant RNA vertebrate viruses, DNA phages, bacteria, and fungi identified (from top to bottom). Numbers in parentheses next to the taxa labels display the ranking in relative abundance for either the BAL or UA metatranscriptome samples, respectively.

**Supplementary Figure 5. Topographical analyses of Metagenome Data.** Comparison of alpha diversity (Shannon Index, **a**) and beta diversity (Bray Curtis Dissimilarity index, **b**) across background negative controls (bronchoscope), bronchoalveolar lavage (BAL) and upper airway (UA) samples (Kruskal-Wallis and PERMANOVA p-values, respectively). (c) Boxplots showing the relative abundance values in log10 across all metagenome samples for the BAL and Upper Airways. The 50 taxa with the highest relative abundance values in the BAL metagenome are displayed; the top 10 in the BAL are highlighted in bold. Each column consists of two plots displaying the most abundant bacteria and fungi identified. Numbers in parentheses next to the taxa labels displays its ranking in relative abundance for either the BAL or UA metagenome samples, respectively.

**Supplementary Figure 6. Evaluation of associations between the lower airway RNA virome and clinical outcome.** Comparisons between the three clinical outcome groups was performed for α diversity (Shannon Index, Kruskal-Wallis p-value, left panel), β diversity (based on Bray Curtis Dissimilarity Index, PERMANOVA p-value, right panel).

**Supplementary Figure 7. Topographical analyses of the bacterial load.** Bacterial load measured by ddPCR targeting 16S rRNA gene in background bronchoscope controls (BKG), lower airway (BAL) and upper airway (UA) samples.

**Supplementary Figure 8. Evaluation of associations between the lower airway mycobiome and clinical outcome.** Fungal taxonomic data was subtracted from metagenome and metatranscriptome data from lower airway samples. **(a)** Comparisons between the three clinical outcome groups was performed for α diversity (Shannon Index, Kruskal-Wallis p-value, left panel), β diversity (based on Bray Curtis Dissimilarity Index, PERMANOVA p-value, right panel) on metagenome data. **(b)** Bubble plot showing DESeq results of fungi enriched in each clinical outcome comparisons based on metagenome data (bubble size based on median relative abundance for those found statistically significant). **(c)** Comparisons between the three clinical outcome groups was performed for α diversity (Shannon Index, Kruskal-Wallis p-value, left panel), β diversity (based on Bray Curtis Dissimilarity Index, PERMANOVA p-value, right panel) on metatranscriptome data. **(d)** Bubble plot showing DESeq results of fungi enriched in each clinical outcome comparisons based on metatranscriptome data (bubble size based on median relative abundance for those found statistically significant).

**Supplementary Figure 9. Evaluation of associations between the lower airway antibiotic resistance genes and clinical outcome.** Bubble plot showing DESeq results of summarized categories of antibiotic resistant microbial genes taken from MEGARes for the metagenome (top) and metatranscriptome (bottom) data sets for each clinical outcome comparison (bubble size based on median relative abundance for those found to be statistically significant). Colored bubbles indicate significantly enriched antibiotic resistance groups.

**Supplementary Figure 10. Measurement of anti-SARS-CoV-2 Immunoglobulin levels and neutralization activity.** Levels of anti-SARS-CoV-2 Spike **(a) and** anti-SARS-CoV-2 receptor binding domain (RBD, **b**) antibodies in BAL from non SARS-CoV-2 infected smoker controls and severely ill COVID-19 intubated patients. Note that the signals for different isotypes cannot be compared because they are detected with different reagents. **(c)** Comparisons of levels of anti-SARS-CoV-2 RBD antibodies in BAL across subjects in different clinical outcome groups (*= Mann–Whitney U p<0.05)**. (d)** Neutralizing activity in BAL samples across subjects in different clinical outcome groups.

**Supplementary Figure 11. Evaluation for associations between the lower airway host tanscriptome and clinical outcome. (a)** PCoA (based on Bray Curtis Dissimilarity Index, PERMANOVA p-value) comparing the three clinical outcome groups. (b, c, d) Volcano plot comparing lower airway host transcriptome between the three clinical outcome groups.

**Supplementary Figure 12. Multi-scale cross-kingdom and co-expression networks. (a)** The neighborhood 5 cross-kingdom metatranscriptome network centered around SARS-CoV-2 is shown. Nodes refer to taxa, edges denote co-abundance after MEGENA. The size of the nodes indicates abundance. Taxa with large nodes are highly abundant. Node-shapes are according to the legend and refer to different microbial kingdoms. The differential abundance of taxa in log2(fold change) between the deceased group and the ≤28-day MV groups is shown by node color - red nodes are taxa abundant in the deceased group compared to the ≤28-day MV group, blue colored nodes denote the opposite. **(b)** Modules M175 and M718 of the host transcriptome are shown. The node size refers to the absolute gene expression value. Nodes with wide node border refer to key regulators/hub genes (see Methods). The differential gene expression of taxa in log2(fold change) between the deceased group and the ≤28-day MV groups is shown by node color - red nodes are up-regulated in the deceased group compared to the ≤28-day MV group, blue colored nodes denote the opposite.

