## Supplementary Figures 1-12 for "Microbial signatures in the lower airways of mechanically ventilated COVID19 patients associated with poor clinical outcome"

### Slide 1
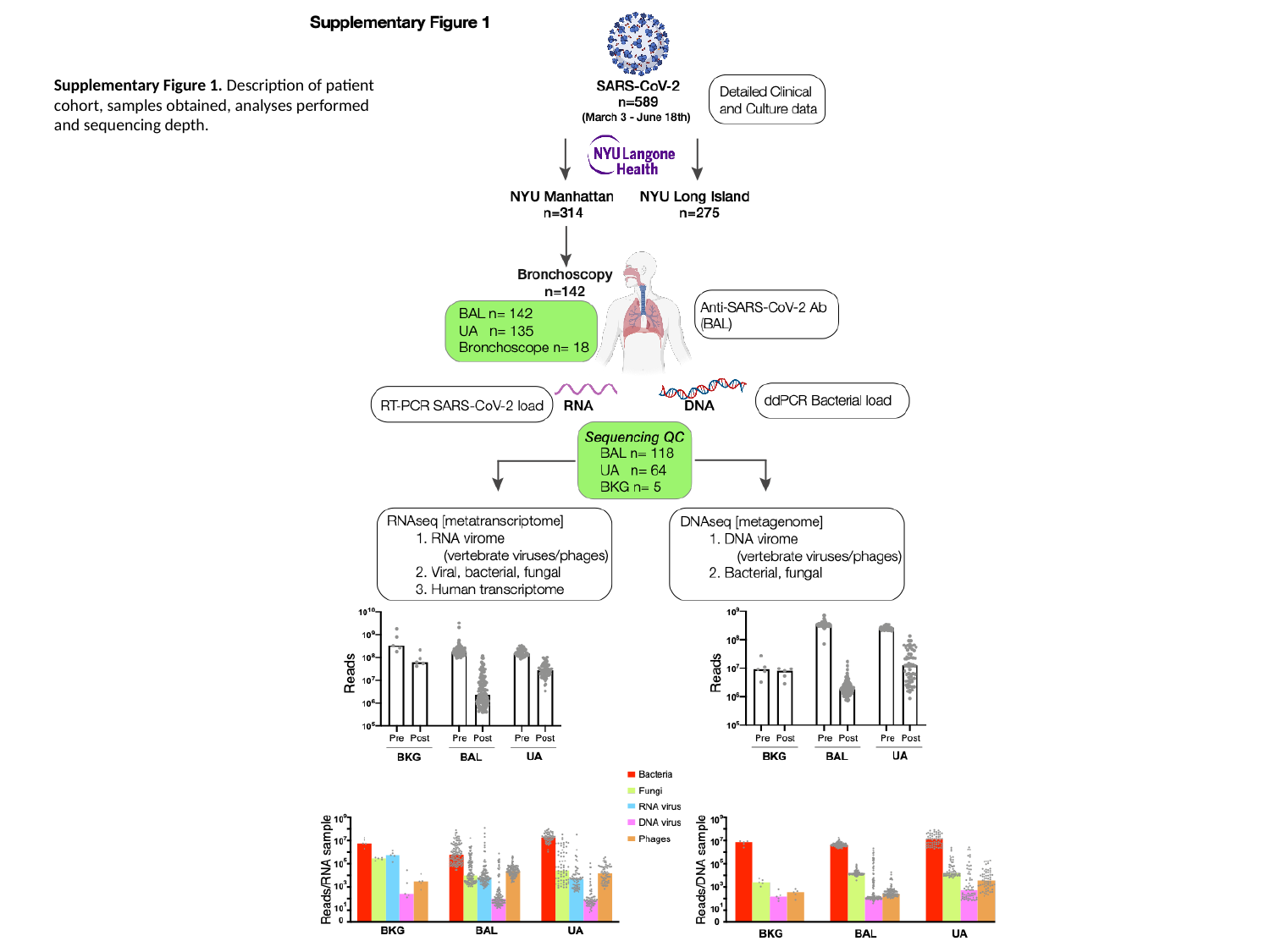

Supplementary Figure 1. Description of patient cohort, samples obtained, analyses performed and sequencing depth.

### Slide 2
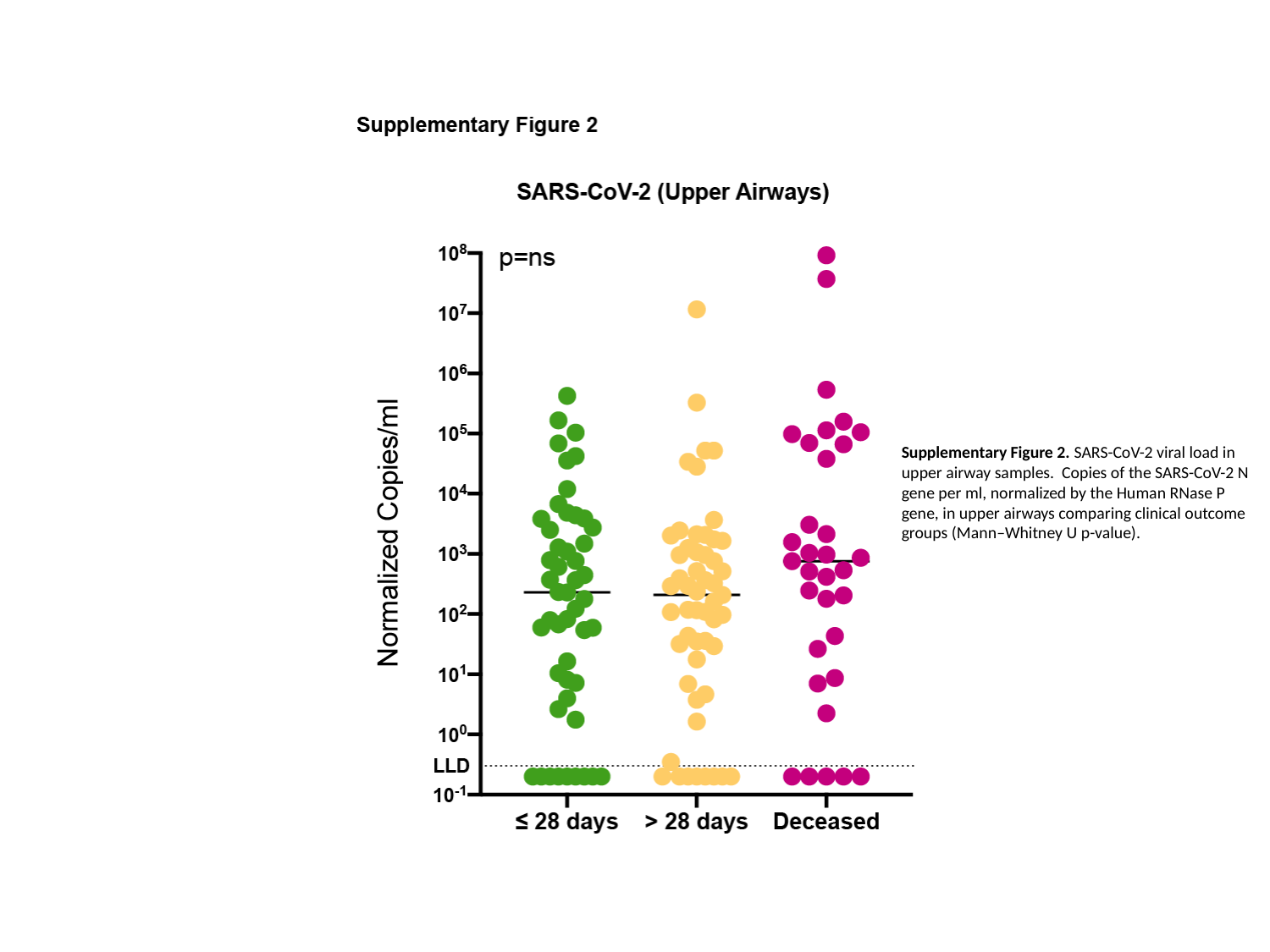

Supplementary Figure 2. SARS-CoV-2 viral load in upper airway samples. Copies of the SARS-CoV-2 N gene per ml, normalized by the Human RNase P gene, in upper airways comparing clinical outcome groups (Mann–Whitney U p-value).

### Slide 3
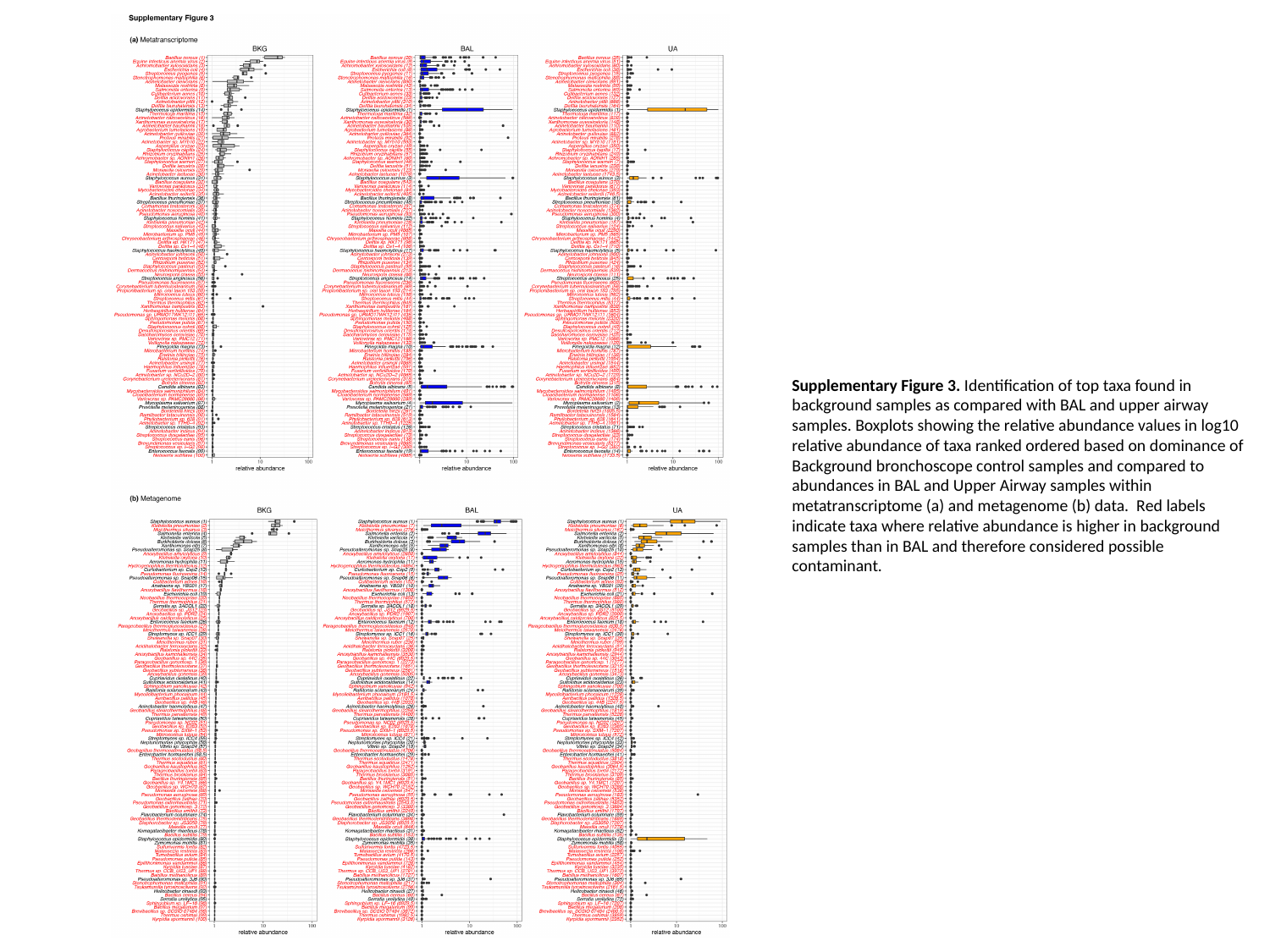

Supplementary Figure 3. Identification of top taxa found in background samples as compared with BAL and upper airway samples. Boxplots showing the relative abundance values in log10 relative abundance of taxa ranked ordered based on dominance of Background bronchoscope control samples and compared to abundances in BAL and Upper Airway samples within metatranscriptome (a) and metagenome (b) data. Red labels indicate taxa where relative abundance is higher in background samples than in BAL and therefore considered possible contaminant.

### Slide 4
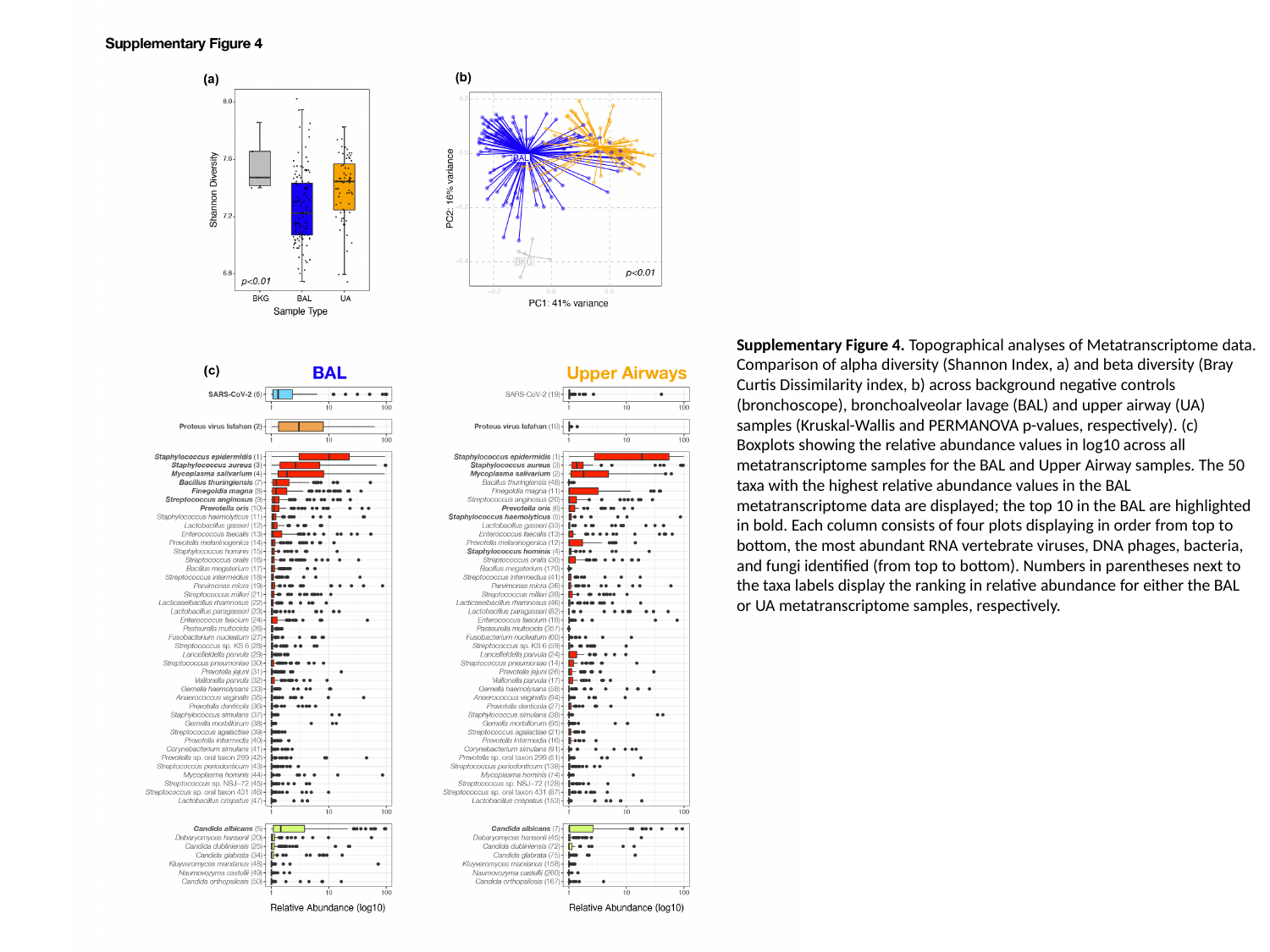

Supplementary Figure 4. Topographical analyses of Metatranscriptome data. Comparison of alpha diversity (Shannon Index, a) and beta diversity (Bray Curtis Dissimilarity index, b) across background negative controls (bronchoscope), bronchoalveolar lavage (BAL) and upper airway (UA) samples (Kruskal-Wallis and PERMANOVA p-values, respectively). (c) Boxplots showing the relative abundance values in log10 across all metatranscriptome samples for the BAL and Upper Airway samples. The 50 taxa with the highest relative abundance values in the BAL metatranscriptome data are displayed; the top 10 in the BAL are highlighted in bold. Each column consists of four plots displaying in order from top to bottom, the most abundant RNA vertebrate viruses, DNA phages, bacteria, and fungi identified (from top to bottom). Numbers in parentheses next to the taxa labels display the ranking in relative abundance for either the BAL or UA metatranscriptome samples, respectively.

### Slide 5
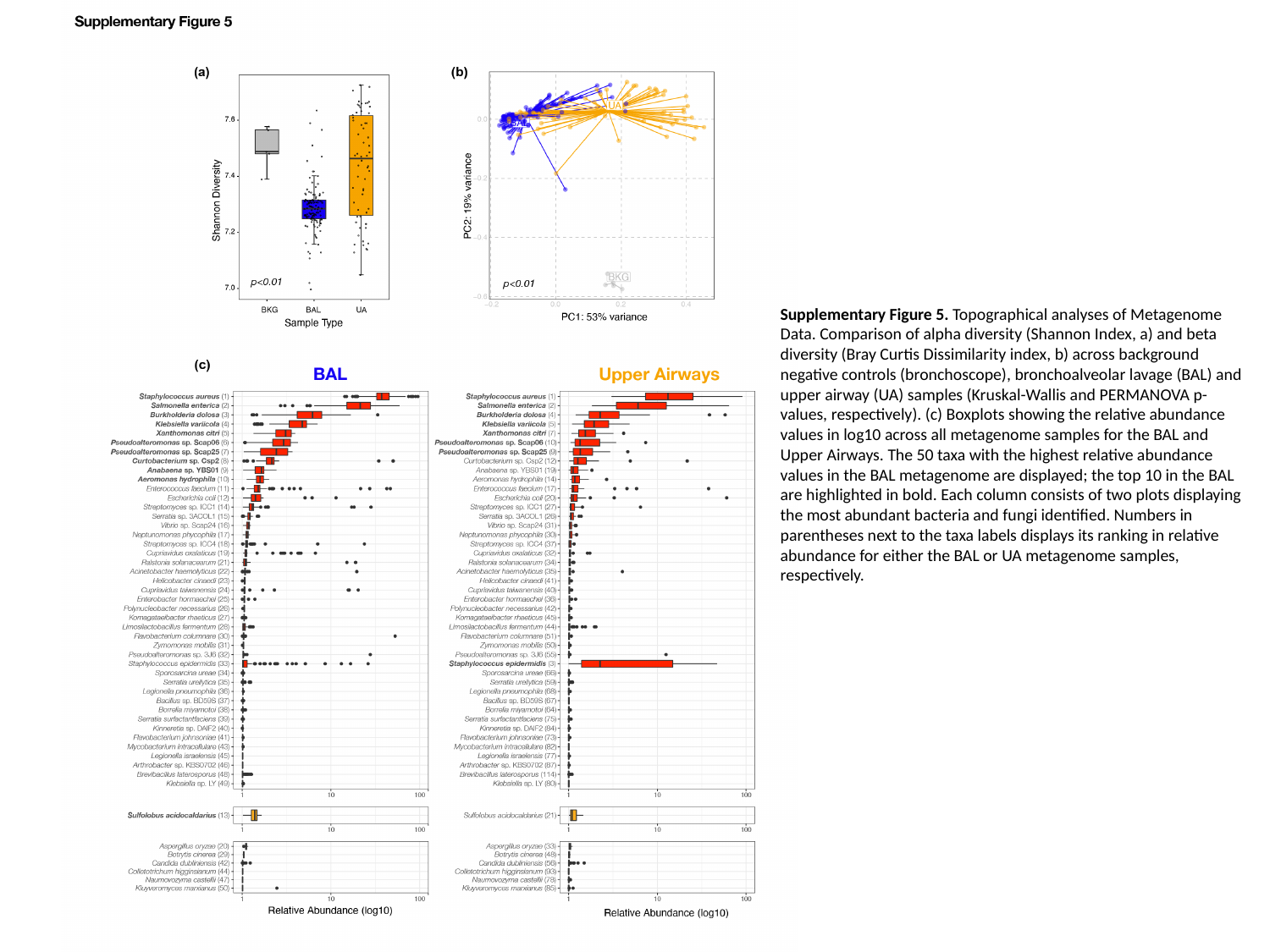

Supplementary Figure 5. Topographical analyses of Metagenome Data. Comparison of alpha diversity (Shannon Index, a) and beta diversity (Bray Curtis Dissimilarity index, b) across background negative controls (bronchoscope), bronchoalveolar lavage (BAL) and upper airway (UA) samples (Kruskal-Wallis and PERMANOVA p-values, respectively). (c) Boxplots showing the relative abundance values in log10 across all metagenome samples for the BAL and Upper Airways. The 50 taxa with the highest relative abundance values in the BAL metagenome are displayed; the top 10 in the BAL are highlighted in bold. Each column consists of two plots displaying the most abundant bacteria and fungi identified. Numbers in parentheses next to the taxa labels displays its ranking in relative abundance for either the BAL or UA metagenome samples, respectively.

### Slide 6
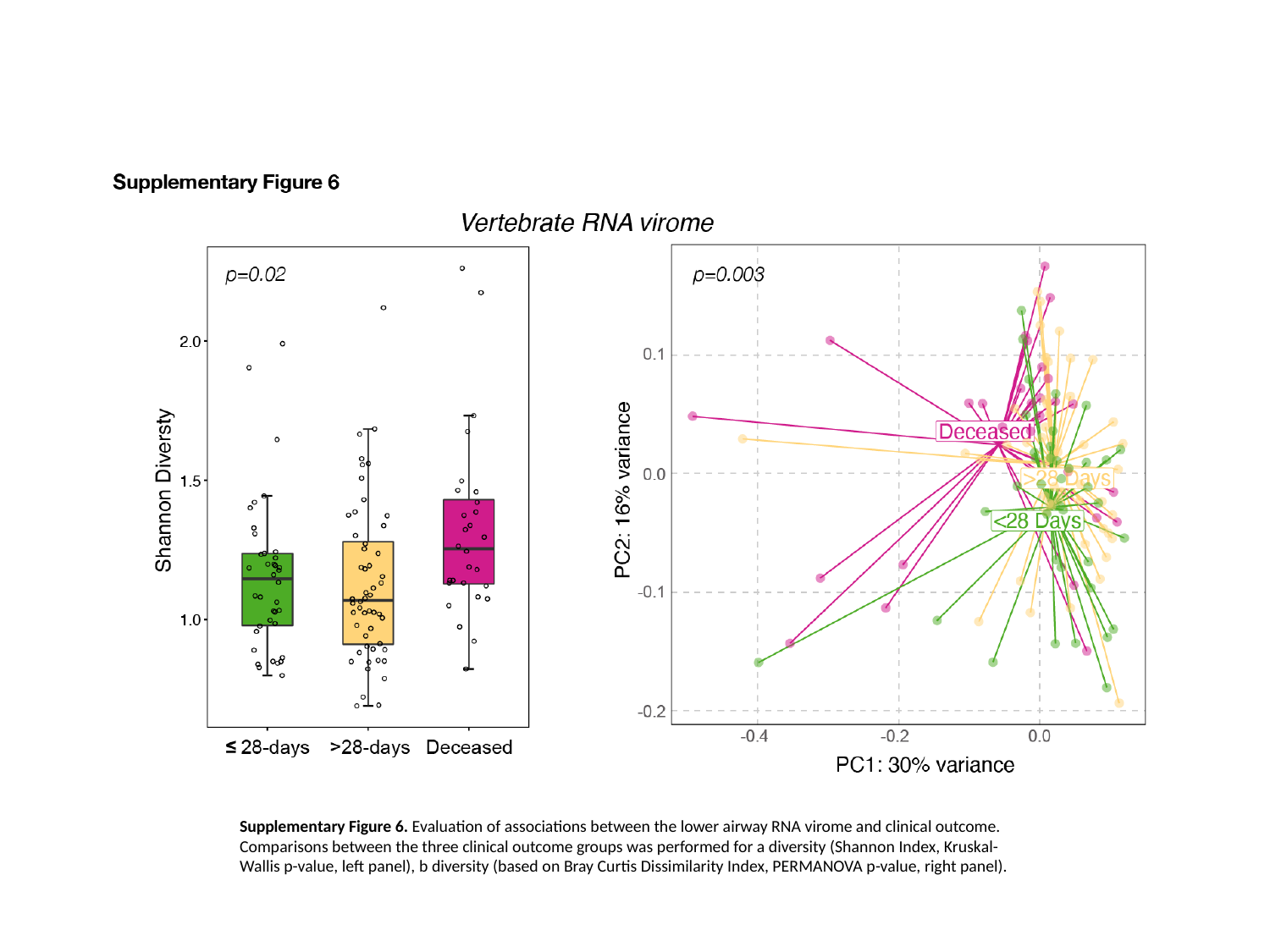

Supplementary Figure 6. Evaluation of associations between the lower airway RNA virome and clinical outcome. Comparisons between the three clinical outcome groups was performed for a diversity (Shannon Index, Kruskal-Wallis p-value, left panel), b diversity (based on Bray Curtis Dissimilarity Index, PERMANOVA p-value, right panel).

### Slide 7
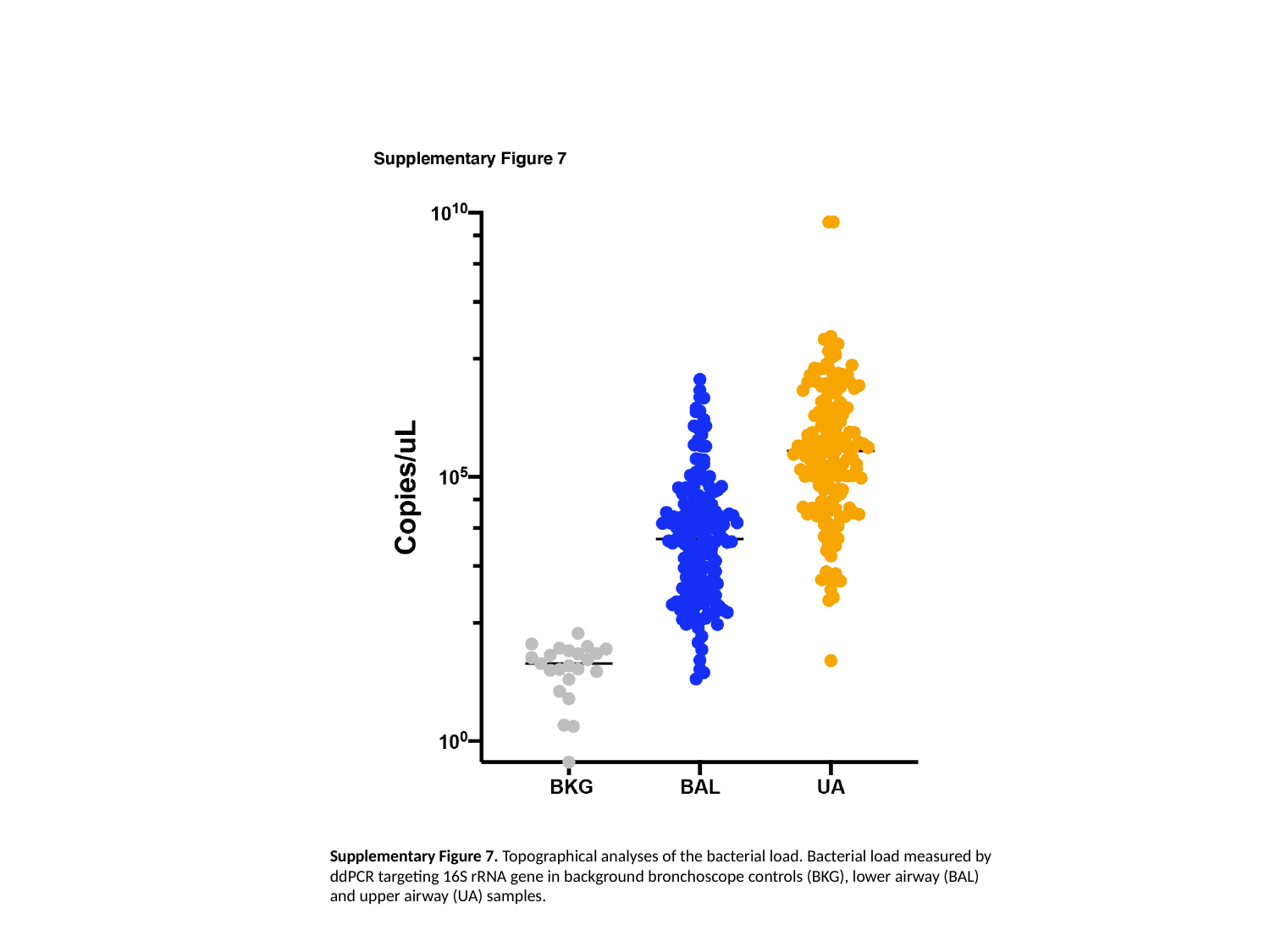

Supplementary Figure 7. Topographical analyses of the bacterial load. Bacterial load measured by ddPCR targeting 16S rRNA gene in background bronchoscope controls (BKG), lower airway (BAL) and upper airway (UA) samples.

### Slide 8
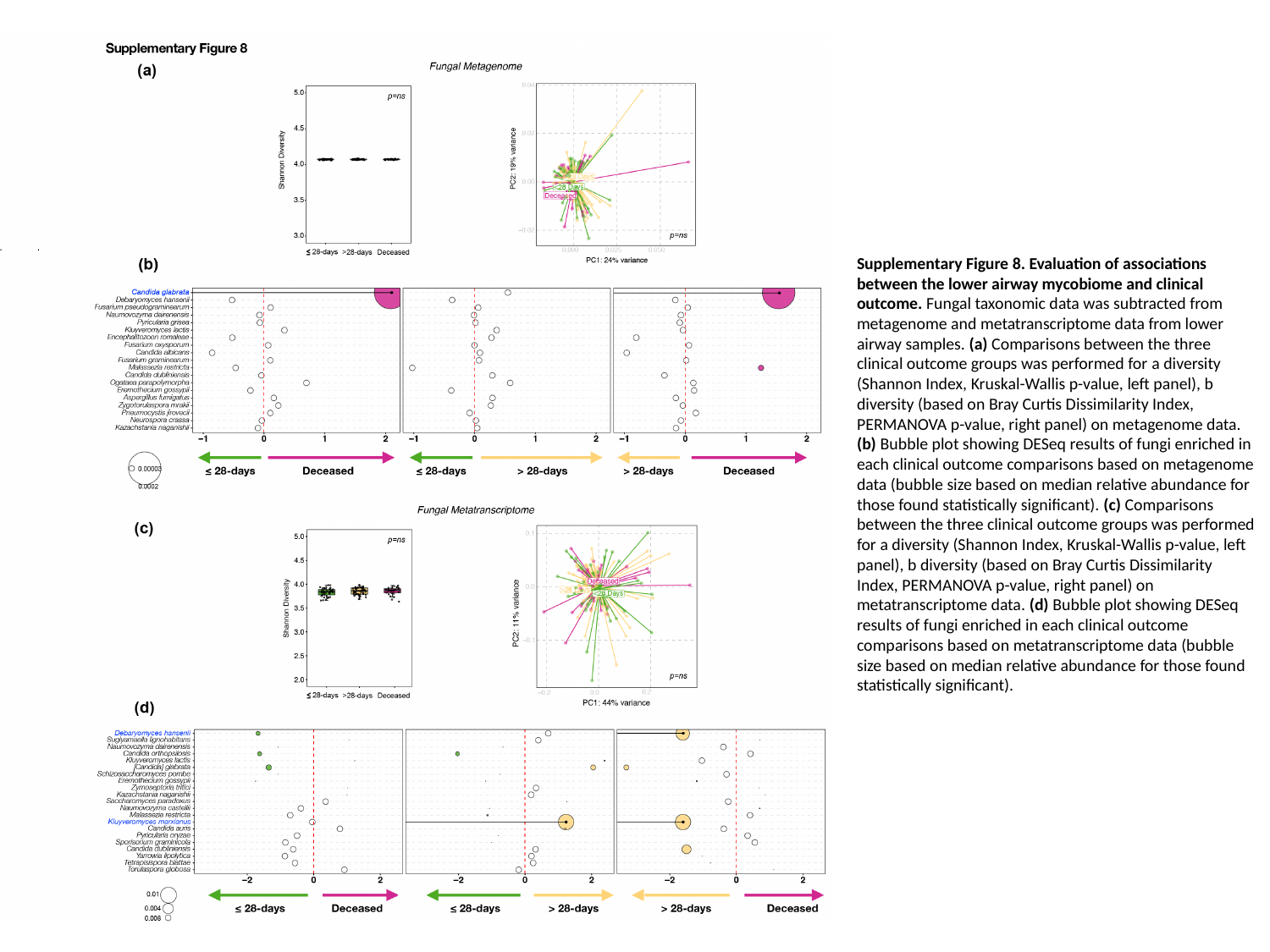

Supplementary Figure 8. Evaluation of associations between the lower airway mycobiome and clinical outcome. Fungal taxonomic data was subtracted from metagenome and metatranscriptome data from lower airway samples. (a) Comparisons between the three clinical outcome groups was performed for a diversity (Shannon Index, Kruskal-Wallis p-value, left panel), b diversity (based on Bray Curtis Dissimilarity Index, PERMANOVA p-value, right panel) on metagenome data. (b) Bubble plot showing DESeq results of fungi enriched in each clinical outcome comparisons based on metagenome data (bubble size based on median relative abundance for those found statistically significant). (c) Comparisons between the three clinical outcome groups was performed for a diversity (Shannon Index, Kruskal-Wallis p-value, left panel), b diversity (based on Bray Curtis Dissimilarity Index, PERMANOVA p-value, right panel) on metatranscriptome data. (d) Bubble plot showing DESeq results of fungi enriched in each clinical outcome comparisons based on metatranscriptome data (bubble size based on median relative abundance for those found statistically significant).

### Slide 9
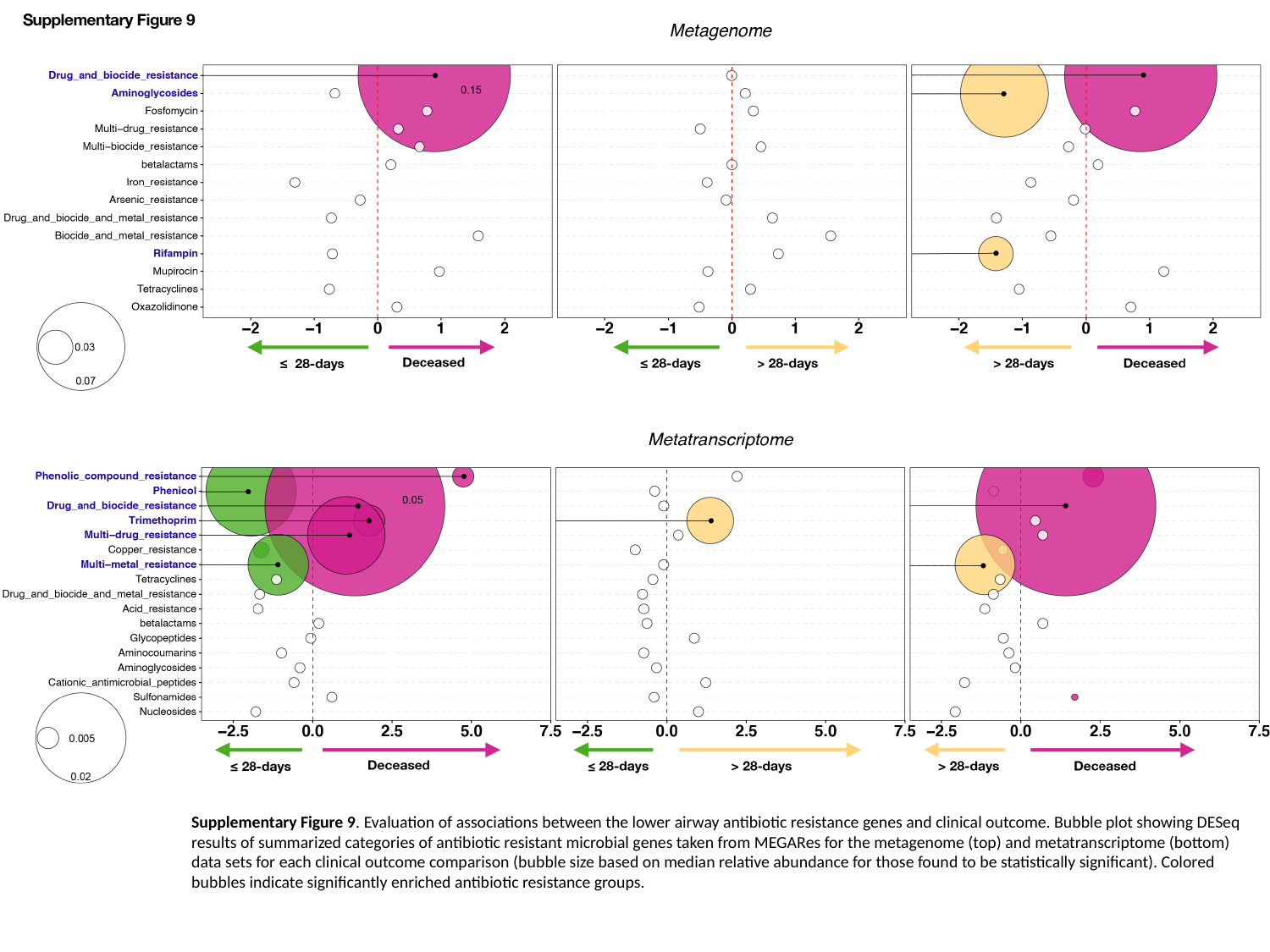

Supplementary Figure 9. Evaluation of associations between the lower airway antibiotic resistance genes and clinical outcome. Bubble plot showing DESeq results of summarized categories of antibiotic resistant microbial genes taken from MEGARes for the metagenome (top) and metatranscriptome (bottom) data sets for each clinical outcome comparison (bubble size based on median relative abundance for those found to be statistically significant). Colored bubbles indicate significantly enriched antibiotic resistance groups.

### Slide 10
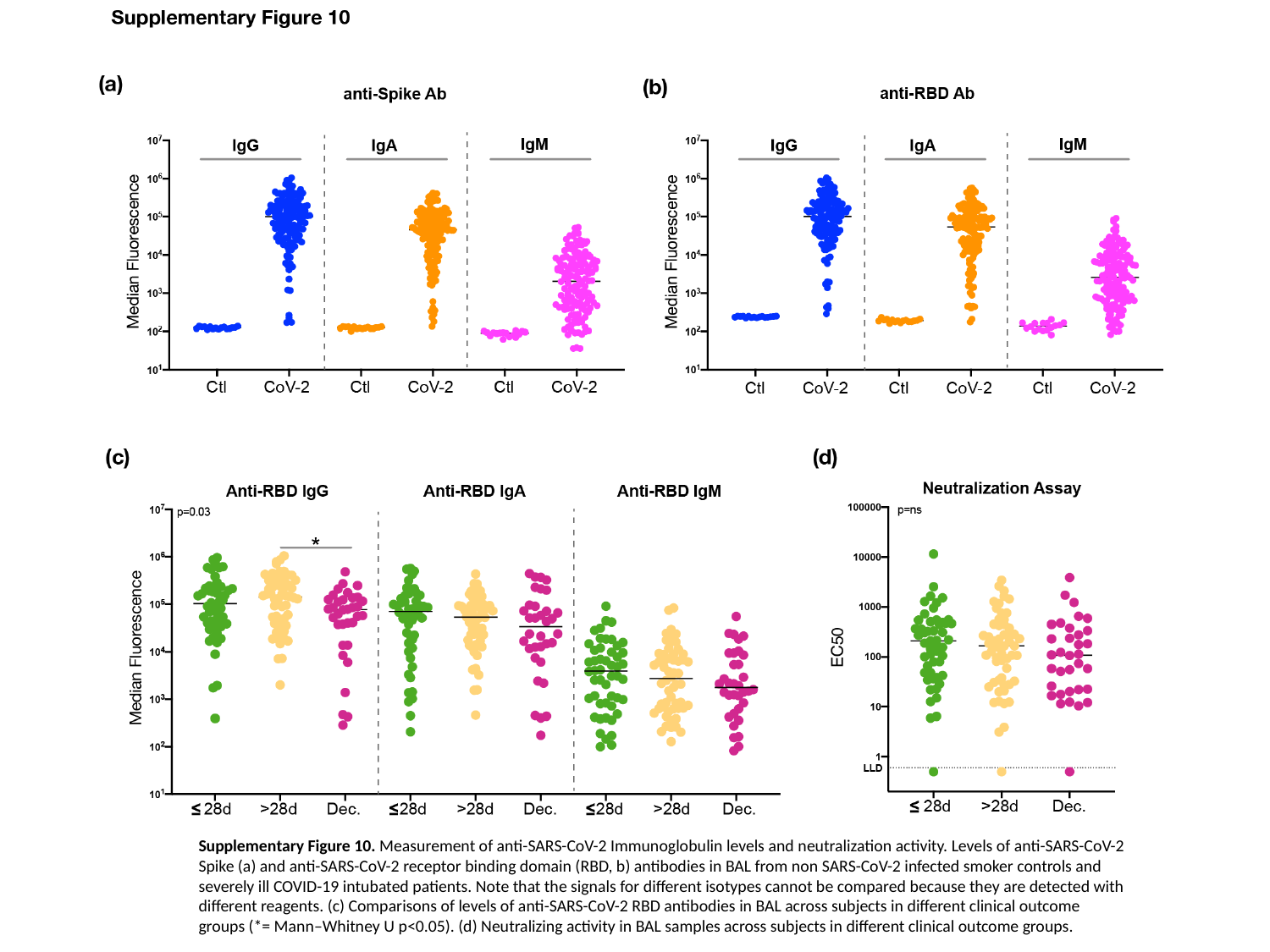

Supplementary Figure 10. Measurement of anti-SARS-CoV-2 Immunoglobulin levels and neutralization activity. Levels of anti-SARS-CoV-2 Spike (a) and anti-SARS-CoV-2 receptor binding domain (RBD, b) antibodies in BAL from non SARS-CoV-2 infected smoker controls and severely ill COVID-19 intubated patients. Note that the signals for different isotypes cannot be compared because they are detected with different reagents. (c) Comparisons of levels of anti-SARS-CoV-2 RBD antibodies in BAL across subjects in different clinical outcome groups (*= Mann–Whitney U p<0.05). (d) Neutralizing activity in BAL samples across subjects in different clinical outcome groups.

### Slide 11
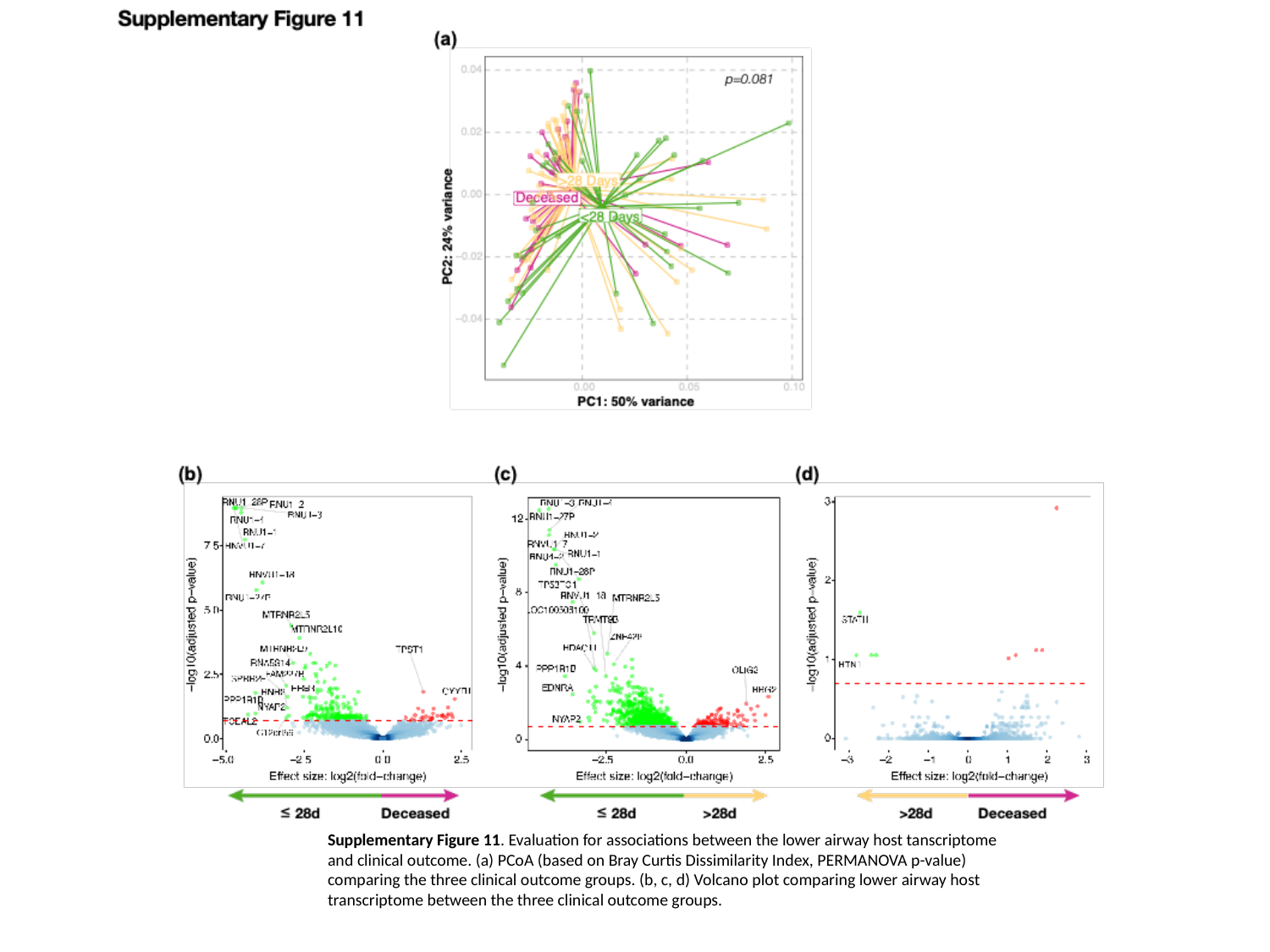

Supplementary Figure 11. Evaluation for associations between the lower airway host tanscriptome and clinical outcome. (a) PCoA (based on Bray Curtis Dissimilarity Index, PERMANOVA p-value) comparing the three clinical outcome groups. (b, c, d) Volcano plot comparing lower airway host transcriptome between the three clinical outcome groups.

### Slide 12
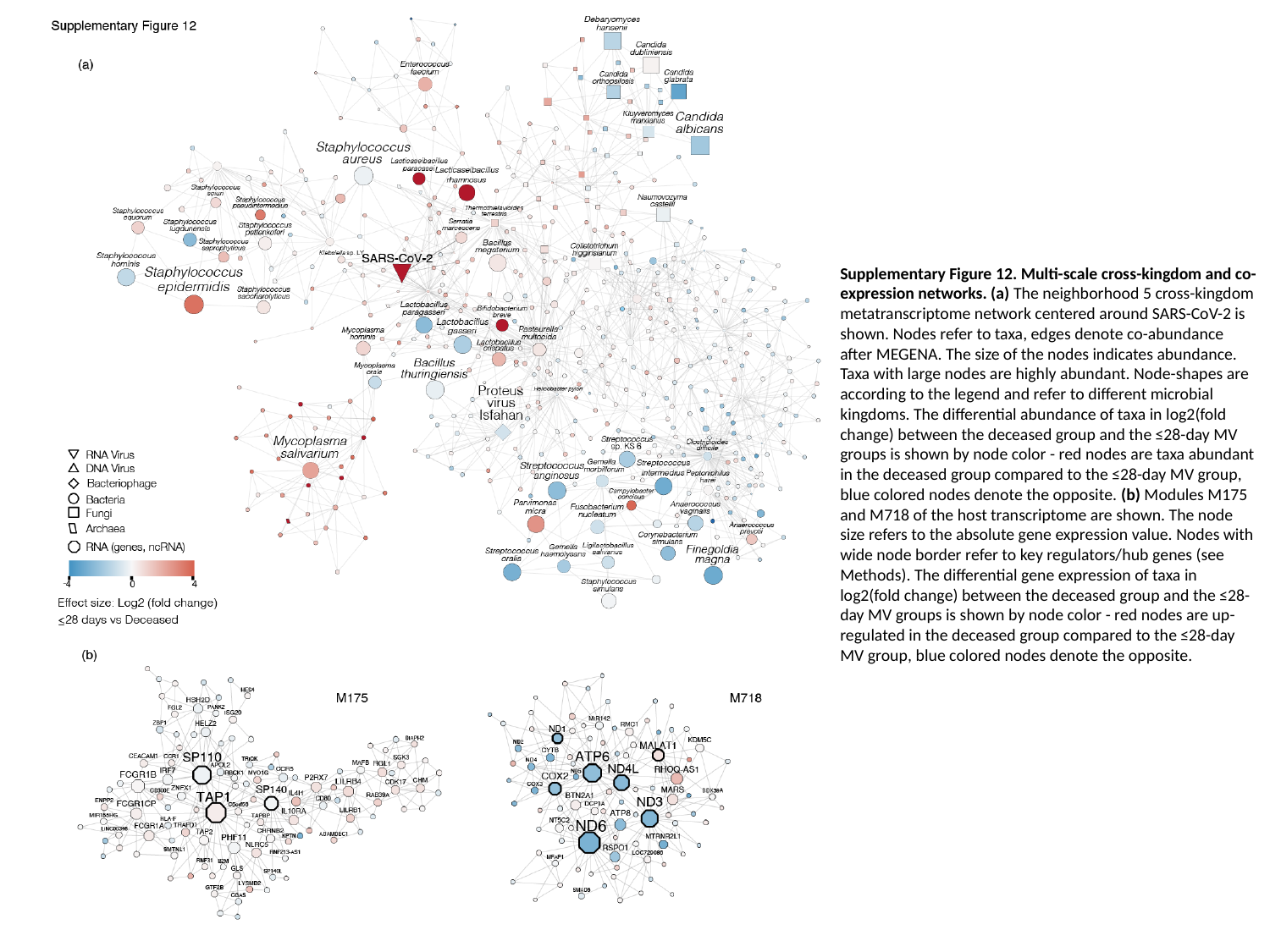

Supplementary Figure 12. Multi-scale cross-kingdom and co-expression networks. (a) The neighborhood 5 cross-kingdom metatranscriptome network centered around SARS-CoV-2 is shown. Nodes refer to taxa, edges denote co-abundance after MEGENA. The size of the nodes indicates abundance. Taxa with large nodes are highly abundant. Node-shapes are according to the legend and refer to different microbial kingdoms. The differential abundance of taxa in log2(fold change) between the deceased group and the ≤28-day MV groups is shown by node color - red nodes are taxa abundant in the deceased group compared to the ≤28-day MV group, blue colored nodes denote the opposite. (b) Modules M175 and M718 of the host transcriptome are shown. The node size refers to the absolute gene expression value. Nodes with wide node border refer to key regulators/hub genes (see Methods). The differential gene expression of taxa in log2(fold change) between the deceased group and the ≤28-day MV groups is shown by node color - red nodes are up-regulated in the deceased group compared to the ≤28-day MV group, blue colored nodes denote the opposite.
